# *Anopheles* salivary antigens as serological biomarkers of vector exposure and malaria transmission: A systematic review with multilevel modelling

**DOI:** 10.1101/2021.09.14.21263589

**Authors:** Ellen A Kearney, Paul A Agius, Victor Chaumeau, Julia C Cutts, Julie A Simpson, Freya JI Fowkes

## Abstract

**Background:** Entomological surveillance for malaria is inherently resource-intensive and produces crude population-level measures of vector exposure which are insensitive in low-transmission settings. Antibodies against *Anopheles* salivary proteins measured at the individual-level may serve as proxy biomarkers for vector exposure and malaria transmission, but their relationship is yet to be quantified.

**Methods:** A systematic review of studies measuring antibodies against *Anopheles* salivary antigens (PROSPERO: CRD42020185449). Multilevel modelling estimated associations between seroprevalence with *Anopheles* human biting rate (HBR) and malaria transmission measures.

**Results:** From 3981 studies identified in literature searches, 42 studies across 16 countries were included contributing 393 meta-observations of anti-*Anopheles* salivary antibodies determined in 42,764 samples. A positive non-linear association between HBR and seroprevalence was found; overall a 50% increase in HBR was associated with a 13% increase in odds of seropositivity (OR: 1.13, 95%CI: 1.06-1.20, *p*<0.001). The association between HBR and *Anopheles* salivary antibodies was strongest with concordant, rather than discordant *Anopheles* species. Seroprevalence was also significantly positively associated with established epidemiological measures of malaria transmission: entomological inoculation rate, *Plasmodium* spp. prevalence, and malarial endemicity class.

**Conclusions:** *Anopheles* salivary antibody biomarkers can serve as a proxy measure for HBR and malaria transmission, and could monitor vectorial capacity and malaria receptivity of a population to sustain malaria transmission. Validation of *Anopheles* species-specific biomarkers are important given the global heterogeneity in the distribution of *Anopheles* species. Salivary biomarkers have the potential to transform surveillance by replacing impractical, inaccurate entomological investigations, especially in areas progressing towards malaria elimination.

**Funding:** Australian National Health and Medical Research Council, Wellcome Trust.

## Introduction

Sensitive and accurate tools to measure and monitor changes in malaria transmission are essential to track progress towards malaria control and elimination goals. Currently, the gold standard measurement of malaria transmission intensity is the entomological inoculation rate (EIR), a population-measure defined as the number of infective *Anopheles* mosquito bites a person receives per unit of time. EIR is calculated as the human biting rate (HBR; measured at the population-level by entomological vector-sampling methodologies (gold standard: human landing catch)) multiplied by the sporozoite index (proportion of captured *Anopheles* with sporozoites present in their salivary glands). However, estimation of EIR and HBR via entomological investigations are inherently labour and resource intensive, requiring trained collectors, specialised laboratories and skilled entomologists. Furthermore, these approaches provide a crude population-level measure of total vector exposure at a particular time and location, precluding investigation of heterogeneity and natural transmission dynamics of individual-level vector-human interactions [1]. For example, indoor human landing catches provide poor estimates of outdoor biting and thus total vector exposure [2]. The sensitivity of EIR is further compromised in low transmission settings where the number of *Plasmodium-*infected specimens detected is low and often zero.

Evaluation of the human antibody response to *Anopheles* spp. salivary proteins has the potential to be a logistically practical approach to estimate levels of exposure to vector bites at an individual-level. Several *Anopheles* salivary proteins have been shown to be immunogenic in individuals naturally exposed to the bites of *Anopheles* vectors and have been investigated as serological biomarkers to measure *Anopheles* exposure [3–11], malaria transmission [12–14] and as an outcome for vector control intervention studies [4-6, 14, 15]. However, a major short-coming of the literature is that studies are largely descriptive and do not quantify the association between entomological and malariometric measures and anti-*Anopheles* salivary antibody responses. We undertook a systematic review with multilevel modelling, to quantify the association between HBR, EIR, and other markers of malaria transmission, with anti-*Anopheles* salivary antibody responses and to understand how these associations vary according to transmission setting and dominant *Anopheles* vectors. This knowledge is pertinent to advance the use of salivary antibody biomarkers as a vector and malaria transmission sero-surveillance tool.

## Methodology

### Search strategy and selection criteria

We performed a systematic review with multilevel modelling according to the MOOSE and PRISMA guidelines [16, 17] (Reporting Standards Document). Five databases were searched for published studies investigating antibodies to *Anopheles* salivary antigens as a biomarker for mosquito exposure or malaria transmission published before 30^th^ of June 2020. The protocol (Supplementary File 1) was registered with PROSPERO (CRD42020185449).

The primary criteria for inclusion in this systematic review was the reporting of estimates of seroprevalence or total levels of Immunoglobulin (Ig) in human sera against *Anopheles* salivary antigens. We considered for inclusion: cross-sectional, cohort, intervention and case-control studies of individuals or populations living in all geographies with natural exposure to *Anopheles* mosquitoes. Studies that were solely performed in participants not representative of the wider naturally exposed population (*i.e.* mosquito allergic patients, soldiers, returned travellers) were excluded.

### Measures

#### Outcomes

The primary outcome of our systematic review was antibodies (seroprevalence or levels, including all Ig isotypes and subclasses) against any *Anopheles* salivary antigens (full-length recombinant proteins, peptides and crude salivary extract). As measurement of antibody levels does not produce a common metric between studies only values of seroprevalence could be included in multilevel modelling analyses. Therefore, to maximise data, authors of studies that reported only antibody levels were contacted and asked to classify their participants as ‘responders’ or ‘non-responders’ according to seropositivity (antibody level relative to unexposed sera). Studies that provided antibody levels or categorised seropositivity based upon arbitrary cut offs are included in narrative terms only.

#### Exposures

The primary exposures of interest were the entomological metrics HBR (average number of bites received per person per night) and EIR (infectious bites received per person per year). Secondary exposures included study-reported prevalence of *Plasmodium* spp. infection (confirmed by either microscopy, rapid diagnostic test, or polymerase chain reaction (PCR)) and seroprevalence of antimalarial antibodies against pre-erythrocytic and blood-stage *Plasmodium* spp. antigens. Where exposure estimates were not provided, we attempted to source data from other publications by the authors, or using the site geolocation and year to obtain estimates of EIR from the Pangaea dataset [18], *P. falciparum* rates in 2-10 year olds (*Pf*PR_2-10_) and dominant vector species (DVS) from the Malaria Atlas Project (MAP) [19]. Malarial endemicity classes were derived by applying established endemicity cut-offs to *Pf*PR_2-10_ estimates [20]. For the purposes of the modelling analyses we defined DVS as where *An. gambiae sensu lato* (*s.l.*) was the only DVS, where *An. gambiae s.l.,* was present with additional DVS, or where *An. gambiae s.l.* was absent. Studies of salivary antigens where exposure variables could not be sourced and data could not be extracted were excluded.

### Statistical analysis

Where seroprevalence estimates of antibodies against the same salivary antigen and exposure of interest were reported in more than one study, generalised multilevel (mixed-effects, logistic) modelling was used to quantify associations between the exposures of interest and salivary antibody seroprevalence measurements [21]. Random intercepts for study and country were included to account for nested dependencies induced from multiple meta-observations (level one) from the same study (level two) and multiple studies from the same country (level three). Additionally, a random slope for the entomological and malariometric exposure parameters was included to model study-specific heterogeneity in the effect of the exposure of interest (HBR/EIR/malaria prevalence/antimalarial antibody seroprevalence). The associations between the various exposures and the different salivary antigens were analysed separately, however estimates of IgG seroprevalence against the recombinant full-length protein (gSG6) and synthetic peptide (gSG6-P1, the one peptide determined in all studies utilising peptides) form of the gSG6 antigen were analysed together.

Potential effect modification of the associations between exposures and anti-*Anopheles* salivary antibody responses were explored. In analyses quantifying the associations between HBR, as well as EIR, and seropositivity, we included an interaction term with DVS and for vector collection method (human landing catch or other indirect measures *e.g.* light traps, spray catches, etc.). For the association between *Plasmodium* spp. prevalence and seropositivity, interaction terms with malaria detection methodology (light microscopy or PCR) and malarial species (*P. falciparum* only, or *P. falciparum* and *P. vivax*) were estimated.

The natural log of the exposure measures (HBR, EIR, malaria prevalence and antimalarial antibody seroprevalence) were estimated in modelling to account for their non-linear associations with seroprevalence. Intraclass correlation coefficients (ICCs) were estimated for country- and study-specific heterogeneity using estimated model variance components. All statistical analyses were performed using STATA v15.1.

### Risk of bias in individual studies

Risk of bias was assessed by one reviewer using the Risk of Bias in Prevalence Studies tool [22]. The risk of bias pertains to the reported seroprevalence estimates of anti-*Anopheles* salivary antibodies included in the multilevel modelling.

## Results

Literature searches identified 158 potentially relevant studies, of which 42 studies were included in the systematic review (Figure 1) and are described in Table 1. Briefly, studies were performed in 16 countries mostly in hypo or mesoendemic areas of Africa where *An. gambiae s.l.* is a dominant vector (n=32), with a minority performed in South America (n=4), Asia (n=4), and the Pacific (n=2) where *An. gambiae s.l.* is not found. In total, our review included 393 meta-observations of anti-*Anopheles* salivary antibodies determined from antibody measurements in a total of 42,764 sera samples. The salivary antigen most commonly assessed was *An. gambiae* Salivary Gland 6 (gSG6), as a full-length protein (n=8) and synthetic peptide (*An. gambiae* Salivary Gland 6 Peptide 1; gSG6-P1; n=24). Additional salivary antigens assessed included *An. gambiae* gSG6-P2 (n=3), recombinant cE5 (n=2), g-5’nuc (n=1), and recombinant *An. funestus* fSG6 (n=2) and f-5’nuc (n=1). Seven studies measured antibodies to whole salivary gland extracts from *An. gambiae* (n=4), *An. darlingi* (n=2), *An. albimanus* (n=1), and *An. dirus* (n=1), while one study assessed antibodies against synthetic peptides of *An. albimanus* (Table 1). All studies investigated total IgG and only five determined an additional isotype or subclass [7, 23–26]. The paucity of studies investigating these latter-mentioned antibody types and *Anopheles* salivary biomarkers precluded extensive multilevel analyses; instead, we present their associations in narrative terms in Supplementary File 5. Analyses reported below focus on quantifying the relationships between HBR, EIR and markers of malaria transmission with total IgG to *An. gambiae* gSG6. The distributions of exposure estimates were: HBR (median: 3.1 bites per person per night, IQR: 0.9-12.1; range: 0-121.4), EIR (median: 7.3 infectious bites received per person per year, IQR: 0-36.4; range: 0-585.6), and *Plasmodium* spp. prevalence (median: 9.1%; IQR: 4-22%; range: 0-94.6%).

**Figure 1.**
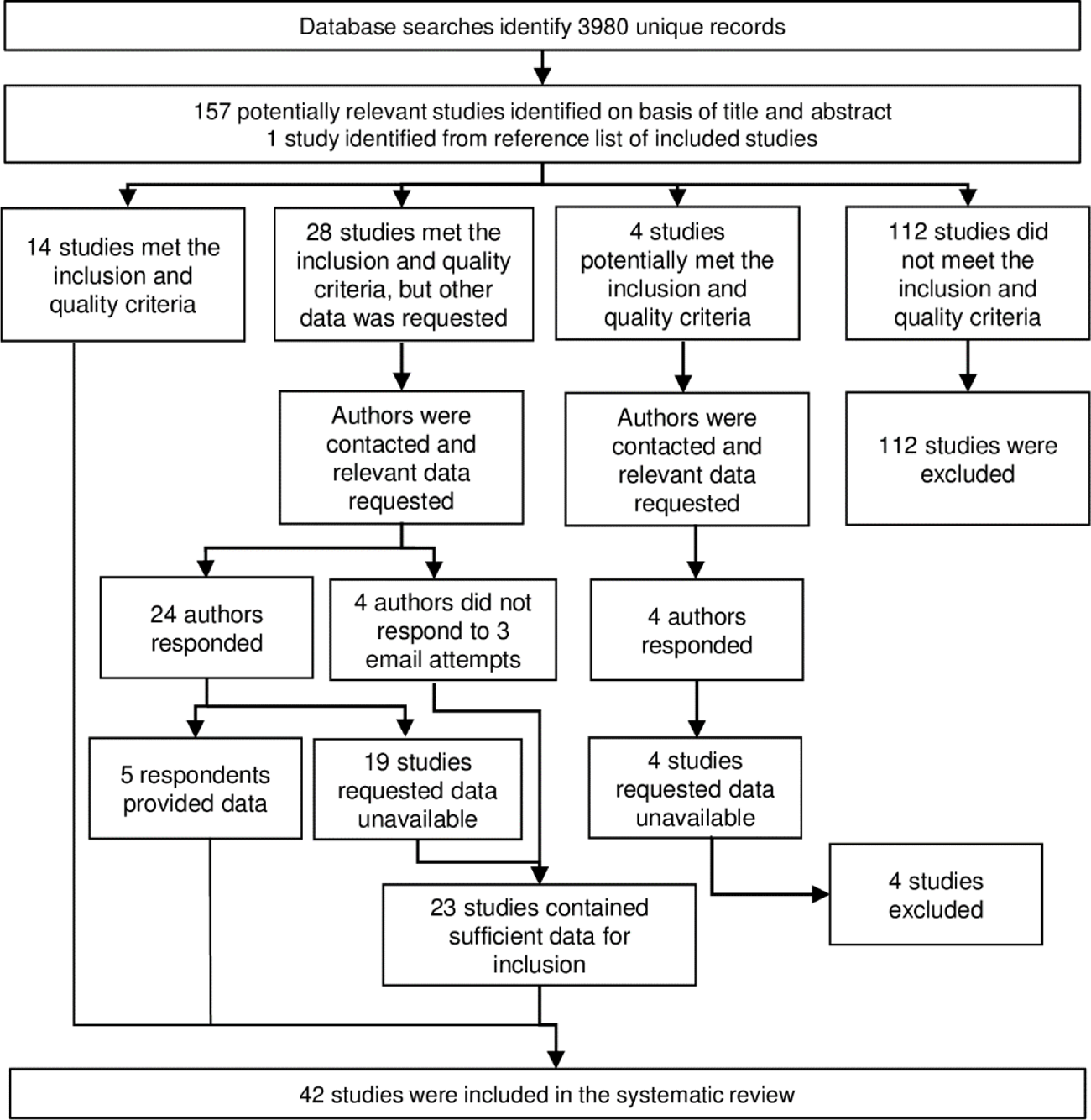
Flow diagram of study identification. Excluded studies are detailed in Figure 1 – Supplement 1.

**Table 1:** Key descriptive information from included studies

| Study year | Country | Malarial endemicity class | Dominant malaria vector species | Study design | No. participants (samples) | Meta-N | Vector and malarionetric variables | Salivary antibody outcomes (Seroprevalence[%];[L]levels) |
| --- | --- | --- | --- | --- | --- | --- | --- | --- |
| **Africa** |  |  |  |  |  |  |  |  |
| Brousseau 2012 [27] | Angola | Hypoendemic; Mesoendemic | *An. funestus* | Cross-sectional<sup>‡</sup> | (1584) | 6 | *Plas*<sup>LM</sup>; *Pf*PR | gSGE IgG [L] |
| Drame 2010 [5] | Angola | Hypoendemic | *An. gambiae s.l.* | Cohort | 105 (1470) | 12 | HBR; *Plas*<sup>LM</sup>; *Pf*PR | gSG6-P1 IgG [%;L] |
| Drame 2010 [6] | Angola | Hypoendemic | *An. gambiae s.l.* | Cohort | 109 (1279) | 12 | HBR; *Plas*<sup>LM</sup>; *Pf*PR | gSGE IgG [L] |
| Marie 2015 [28] | Angola | Hypoendemic | *An. gambiae s.l.* | Cohort | 71 (852) | 12 | HBR; *Pf*PR | gcE5 IgG [L] |
| Drame 2015 [7] | Benin | Hyperendemic | *An. gambiae s.l.*; *An. funestus* | Cohort<sup>‡</sup> | 133 (532) | 4 | HBR; *Pf*PR | gSG6-P1 IgG & IgM [%;L] |
| Rizzo 2011 [9] | Burkina Faso | Hyperendemic* | *An. gambiae s.l.* | Repeated cross-sectional | (2066) | 14 | HBR; EIR; *Plas*<sup>LM§</sup> | gSG6 IgG [%;L] |
| Rizzo 2011 [8] | Burkina Faso | Hyperendemic* | *An. gambiae s.l.* | Repeated cross-sectional | 335 (335) | 3 | HBR | fSG6 IgG [%;L] |
| Rizzo 2014 [24] | Burkina Faso | Hyperendemic* | *An. gambiae s.l.* | Repeated cross-sectional | (359) | 3 | HBR | gcE5 IgG [%;L]; IgG1 & IgG4 [L] |
| Rizzo 2014 [25] | Burkina Faso | Hyperendemic* | *An. gambiae s.l.* | Repeated cross-sectional | 270 (270) | 6 | HBR | gSG6 IgG1 & IgG4 [L] |
| Soma 2018 [29] | Burkina Faso | Mesoendemic | *An. gambiae s.l.* | Cross-sectional | 1728 (273) | 6 | HBR; EIR; *Plas*<sup>LM</sup>; *Pf*PR | gSG6-P1 IgG [%;L] |
| Koffi 2015 [30] | Cote d'Ivoire | Hypoendemic; Mesoendemic | *An. gambiae s.l.*; *An. funestus*<sup>†</sup> | Cross-sectional | 94 (94) | 3 | *Plas*<sup>LM</sup>; *Pf*-IgG; *Pf*PR | gSG6-P1 IgG [%;L] |
| Koffi 2017 [31] | Cote d'Ivoire | Hypoendemic | *An. gambiae s.l.*; *An. funestus*<sup>†</sup> | Repeated cross-sectional | 234 (234) | 5 | *Pf*-IgG; *Pf*PR | gSG6-P1 IgG [%;L] |
| Traoré 2018 [32] | Cote d'Ivoire | Hypoendemic | *An. gambiae s.l.*; *An. funestus*<sup>†</sup> | Repeated cross-sectional<sup>‡</sup> | 89 (178) | 4 | HBR; *Plas*<sup>LM</sup>; *Pf*PR | gSG6-P1 IgG [L] |
| Traoré 2019 [33] | Cote d'Ivoire | Hypoendemic | *An. gambiae s.l.*; *An. funestus*<sup>†</sup> | Repeated cross-sectional<sup>‡</sup> | (442) | 6 | HBR; *Plas*<sup>LM</sup>; *Pf*PR | gSG6-P1 IgG [%;L] |
| Sadia-Kacou 2019 [34] | Cote d'Ivoire | Mesoendemic | *An. gambiae s.l.* | Repeated cross-sectional<sup>‡</sup> | 775 (775) | 8 | *PfPR* | *gSG6-P1 IgG [L]* |
| Badu 2015 [35] | Ghana | Mesoendemic | *An. gambiae s.l.*; *An. funestus*<sup>†</sup> | Repeated cross-sectional<sup>‡</sup> | 295 (885) | 3 | *Plas*<sup>LM</sup>; *Pf-IgG*; *PfPR* | *gSG6-P1 IgG [%;L]* |
| Badu 2012 [3] | Kenya | Hypoendemic; Mesoendemic | *An. gambiae s.l.* | Repeated cross-sectional | (1366) | 5 | EIR; *Plas*<sup>LM</sup>; *PfPR* | *gSG6-P1 IgG [%;L]* |
| Sagna 2013[36] | Senegal | Hypoendemic; Mesoendemic | *An. gambiae s.l.* | Cohort<sup>‡</sup> | 265 (1325) | 25 | HBR; *Plas*<sup>LM</sup>; *PfPR* | *gSG6-P1 IgG [%;L]* |
| Drame 2012 [11] | Senegal | Hypoendemic | *An. gambiae s.l.* | Cross-sectional | 1010 (1010) | 16 | HBR; *PfPR* | *gSG6-P1 IgG [%;L]* |
| Poinsignon 2010 [37] | Senegal | Hypoendemic | *An. funestus* | Cohort<sup>‡</sup> | 87 (261) | 3 | HBR; *Plas*<sup>LM</sup>; *PfPR* | *gSG6-P1 IgG [L]* |
| Sarr 2012 [38] | Senegal | Hypoendemic; Mesoendemic | *An. gambiae s.l.*; *An. funestus*<sup>†</sup> | Repeated cross-sectional<sup>‡</sup> | (401) | 4 | HBR; *Plas*<sup>LM</sup>; *Pf-IgG*; *PfPR* | *gSG6-P1 IgG [%;L]* |
| Lawaly 2012 [23] | Senegal | Mesoendemic | *An. gambiae s.l.*; *An. funestus*<sup>†</sup> | Cohort | 387 (711) | 4 | HBR; *Plas*<sup>LM</sup>; *PfPR* | *gSGE IgG, IgG4 & IgE [L]* |
| Ali 2012 [39] | Senegal | Hypoendemic;* Mesoendemic;* Hyperendemic* | *An. gambiae s.l.*; *An. funestus*; *An. pharoensis* | Cross-sectional | (134) | 3 | HBR; EIR | *gSG6 IgG [%;L]*<br>*fSG6 IgG [%;L]*<br>*f5'nuc IgG [%;L]*<br>*g5'nuc IgG [%;L]* |
| Ambrosino 2010 [40] | Senegal | Hypoendemic;* Mesoendemic;* Hyperendemic* | *An. gambiae s.l.*; *An. funestus*; *An. pharoensis* | Cross-sectional | (123) | 3 | EIR; *Pf-IgG* | *gSG6-P1 IgG [%]*<br>*gSG6-P2 IgG [%]* |
| Perraut 2017 [41] | Senegal | Hypoendemic; Mesoendemic | *An. gambiae s.l.*; *An. funestus* | Repeated cross-sectional | (798) | 4 | EIR; *Plas*<sup>LM</sup>; *Plas*<sup>PCR</sup>; *Pf-IgG*; *PfPR* | *gSG6-P1 IgG [%]* |
| Poinsignon 2008 [42] | Senegal | Mesoendemic | *An. gambiae s.l.* | Cross-sectional<sup>‡</sup> | 241 (241) | 3 | HBR; *PfPR* | *gSG6-P1 IgG [L]*<br>*gSG6-P2 IgG [L]* |
| Poinsignon 2009 [43] | Senegal | Mesoendemic | *An. gambiae s.l.* | Repeated cross-sectional<sup>‡</sup> | 61 (122) | 2 | HBR; *Plas*<sup>LM</sup>; *PfPR* | *gSG6-P1 IgG [L]* |
| Remoue 2006 [44] | Senegal | Mesoendemic | *An. gambiae s.l.* | Cross-sectional<sup>‡</sup> | 448 (448) | 4 | HBR; *Plas*<sup>LM</sup>; *PfPR* | *gSGE IgG [%;L]* |
| Sagna 2019 [45] | Senegal | Hypoendemic | *An. gambiae s.l.* | Cross-sectional<sup>‡</sup> | 809 (809) | 4 | *PfPR* | *gSG6-P1 IgG [L]* |
| Stone 2012 [10] | Tanzania | Mesoendemic;<br>Hyperendemic | *An. gambiae s.l.* | Cross-sectional<sup>‡</sup> | 636<br>(636) | 16 | HBR; *Pf*-IgG; *Pf*PR | *g*SG6 IgG [%;L] |
| Yman 2016 [46] | Tanzania | Mesoendemic;<br>Holoendemic* | *An. gambiae s.l.*;<br>*An. funestus* | Repeated cross-sectional<sup>‡</sup> | 668<br>(668) | 16 | *Pf*-IgG; *Pf*PR | *g*SG6 IgG [%] |
| Proietti 2013<br>[47] | Uganda | Mesoendemic | *An. gambiae s.l.*;<br>*An. funestus*<sup>†</sup> | Repeated cross-sectional | 509<br>(509) | 3 | *Pf*-IgG; *Pf*PR | *g*SG6 IgG [%] |
| **Americas** |  |  |  |  |  |  |  |  |
| Andrade 2009<br>[48] | Brazil | Eliminating;<br>Hypoendemic | *An. darlingi* | Cross-sectional | 204<br>(204) | 3 | *Plas*+<sup>LM¶</sup>;<br>*Plas*+<sup>PCR¶</sup>; *Pf*PR | *d* SGE IgG [L ] |
| Londono-Renteria 2015<br>[12] | Colombia |  | *An. albimanus* | Cross-sectional | 42<br>(42) | 2 | *Plas*+<sup>PCR¶</sup> | *g* SG6-P1 IgG [L ] |
| Londono-Renteria 2020<br>[49] | Colombia | Eliminating | *An. albimanus* | Cross-sectional | 337<br>(337) | 2 | *Plas*+<sup>PCR</sup>; *Pf*PR | *a*PEROX-P1, P2 & P3 IgG [L];<br>*a*TRANS-P1 & P2 IgG [L] |
| Montiel 2020<br>[50] | Colombia | Eliminating | *An. albimanus* | Case-control | 113<br>(113) | 2 | *Plas*+<sup>LM</sup>; *Plas*+<sup>PCR¶</sup>;<br>*Pf*PR | *g* SG6-P1 IgG [L ];<br>*d* SGE IgG [L ];<br>*a* STECLA SGE IgG [L ];<br>*a* Cartagena SGE IgG [L ] |
| **Asia** |  |  |  |  |  |  |  |  |
| Kerkhof 2016<br>[51] | Cambodia | Hypoendemic | *An. dirus* | Cross-sectional | (8438) | 113 | *Plas*+<sup>PCR</sup>; *Pf*-IgG;<br>*Pv*-IgG; *Pf*PR | *g*SG6-P1 IgG [%;L];<br>*g*SG6-P2 IgG [%;L] |
| Charlwood 2017<br>[52] | Cambodia | Eliminating | *An. dirus* | Repeated cross-sectional | 454<br>(1180) | 6 | HBR; *Pf*-IgG; *Pf*PR | *g*SG6 IgG [L] |
| Ya-Umphun<br>2017 [13] | Myanmar | Eliminating | *An. minimus*;<br>*An. maculatus* | Repeated cross-sectional | 2602<br>(9425) | 28 | HBR; EIR;<br>*Plas*+<sup>PCR</sup>;<br>*Pf*-IgG; *Pf*PR | *g*SG6-P1 IgG [%;L] |
| Waitayakul<br>2006 [26] | Thailand |  | *An. dirus* | Case-control | 139<br>(139) | 3 | *Plas*+<sup>LM</sup> | *dir* SGE IgG & IgM [L ] |
| **Pacific** |  |  |  |  |  |  |  |  |
| Pollard 2019<br>[53] | Solomon<br>Islands | Eliminating;<br>Hypoendemic | *An. farauti* | Repeated cross-sectional | 686<br>(791) | 9 | HBR; EIR; *Pf*PR | *g*SG6-P1 IgG [%;L] |
| Idris 2017 [15] | Vanuatu | Eliminating;<br>Hypoendemic;<br>Mesoendemic | *An. farauti* | Repeated cross-sectional | 905<br>(905) | 3 | *Plas*+<sup>LM</sup>; *Pf*-IgG;<br>*Pv*-IgG; *Pf*PR | *g*SG6 IgG [%;L] |

Generalised multilevel modelling of 132 meta-observations from 12 studies estimated a positive association between *Anopheles* spp.-HBR and seroprevalence of IgG to *An. gambiae* gSG6 salivary antigen [5, 7, 8, 10, 11, 13, 29, 33, 36, 38, 39, 53]. The magnitude of the association was such that a 50% relative increase in HBR was associated with a 13% increase (OR: 1.13; 95%CI: 1.06-1.20, *p*<0.001) in the odds of anti-gSG6 IgG seropositivity (Figure 2 and Figure 2 – Supplement 1). As the association between gSG6 IgG and HBR (log transformed) is non-linear, relative change in gSG6 IgG will vary across HBR levels. To quantify this, we estimated odds ratios for different incremental per cent increases in HBR, shown in Figure 3A. Heterogeneity in the effect of HBR on gSG6 across studies was observed (likelihood ratio χ^2^ (1) = 109.25, *p*<0.001); the 95% reference range of study-specific effects for a 50% relative increase in HBR ranged from a 7% reduction to a 37% increase in odds (OR: 0.93-1.37). There was no evidence (*p*=0.443) that the association between HBR and gSG6 IgG varied according to vector collection method (human landing catch or other indirect methods). However, the magnitude of the association between *An. gambiae* gSG6 IgG seropositivity and HBR was greatest where *An. gambiae s.l.* was the only dominant vector (*p*<0.001); a 50% relative increase in HBR was associated with a 20% increase (OR: 1.20; 95%CI: 1.11-1.31; *p*<0.001) in the odds of gSG6 IgG seropositivity, compared to an 8% (OR: 1.08; 95%CI: 0.99-1.18; *p*=0.079) and 3% (OR: 1.03; 95%CI: 1.02-1.05; *p*<0.001) increase in odds of gSG6 IgG seropositivity where other DVS were present in addition to *An. gambiae s.l.* and where *An. gambiae s.l.* was absent, respectively (Figure 4; Figure 4 – Supplement 1). In order to quantify the relationship between gSG6 IgG seroprevalence and HBR, for given HBR values we estimated gSG6 IgG seroprevalence by producing model-based predicted probabilities overall and by DVS (Figure 4). Where *An. gambiae s.l* is the only DVS, predicted seroprevalence of *An. gambiae* gSG6 ranged from 8% (95%CI: 0-22%) to 86% (95%CI: 67-100%) for an HBR of 0.01 to 100 bites per person per night respectively.

**Figure 2.**
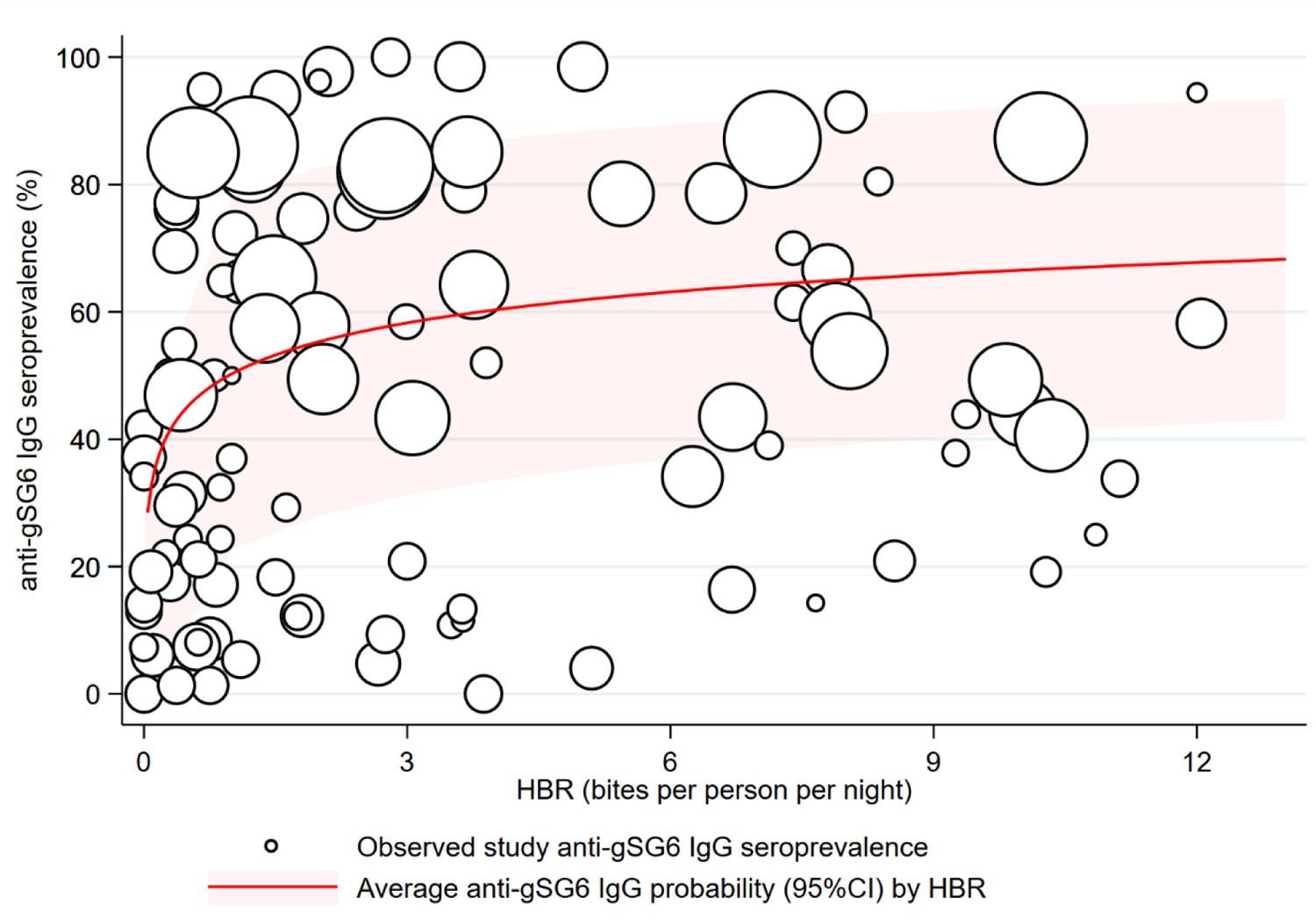
Association between anti-gSG6 IgG seroprevalence and human biting rate (HBR). Figure shows the observed anti-gSG6 IgG (either recombinant or peptide form) seroprevalence (%) and HBR for each meta-observation, as well as the predicted average anti-gSG6 IgG seroprevalence (predicted probability for the average study and country) with 95% confidence intervals (95%CI). Circles are proportional to the size of the sample for each meta-observation. Association estimated using generalised multilevel mixed-effects modelling to account for the hierarchical nature of the data, using an anti-gSG6 IgG meta-observation, nested within study nested in country (model output shown in Figure 2 – Supplement 1; *p*<0.001). Note: to aid visualisation the graph is truncated to show 75% of HBR data (*i.e.* 75^th^ percentile 12.2 bites per person per night) but HBR estimates up to 121.4 were observed and analysed.

**Figure 3.**
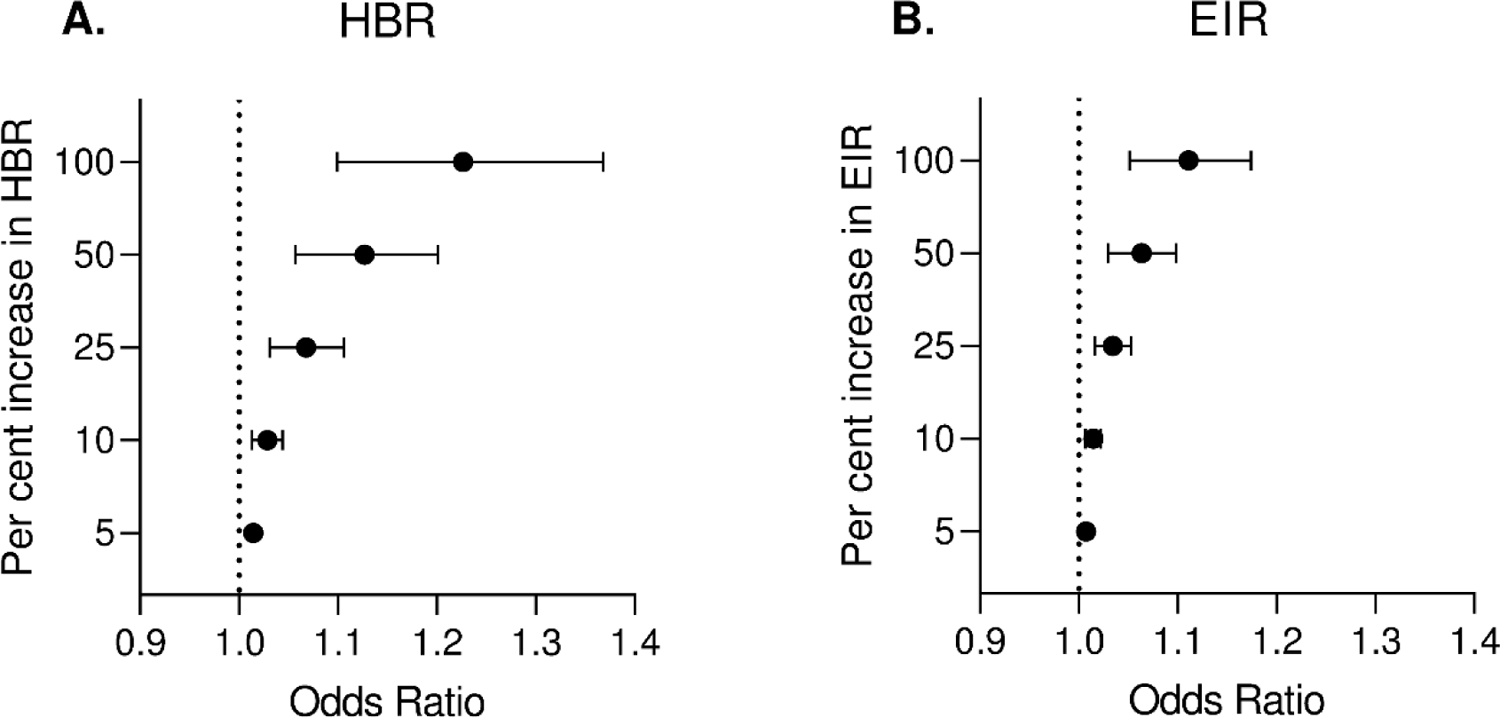
Estimated change in odds of anti-gSG6 IgG seropositivity for given per cent increases in A) HBR (bites/person/night) and B) EIR (infective bites/person/year). Forest plots show estimated odds ratios (ORs) with 95% confidence intervals for given per cent increases in HBR or EIR, estimated using generalised multilevel mixed-effects modelling of the association between anti-gSG6 IgG seropositivity and log HBR or EIR, with random-effects for country-specific and study-specific heterogeneity in gSG6 IgG seroprevalence and study-specific heterogeneity in effect of HBR or EIR (see Figure 2 – Supplement 1 and Figure 3 – Supplement 1 for model output).

**Figure 4.**
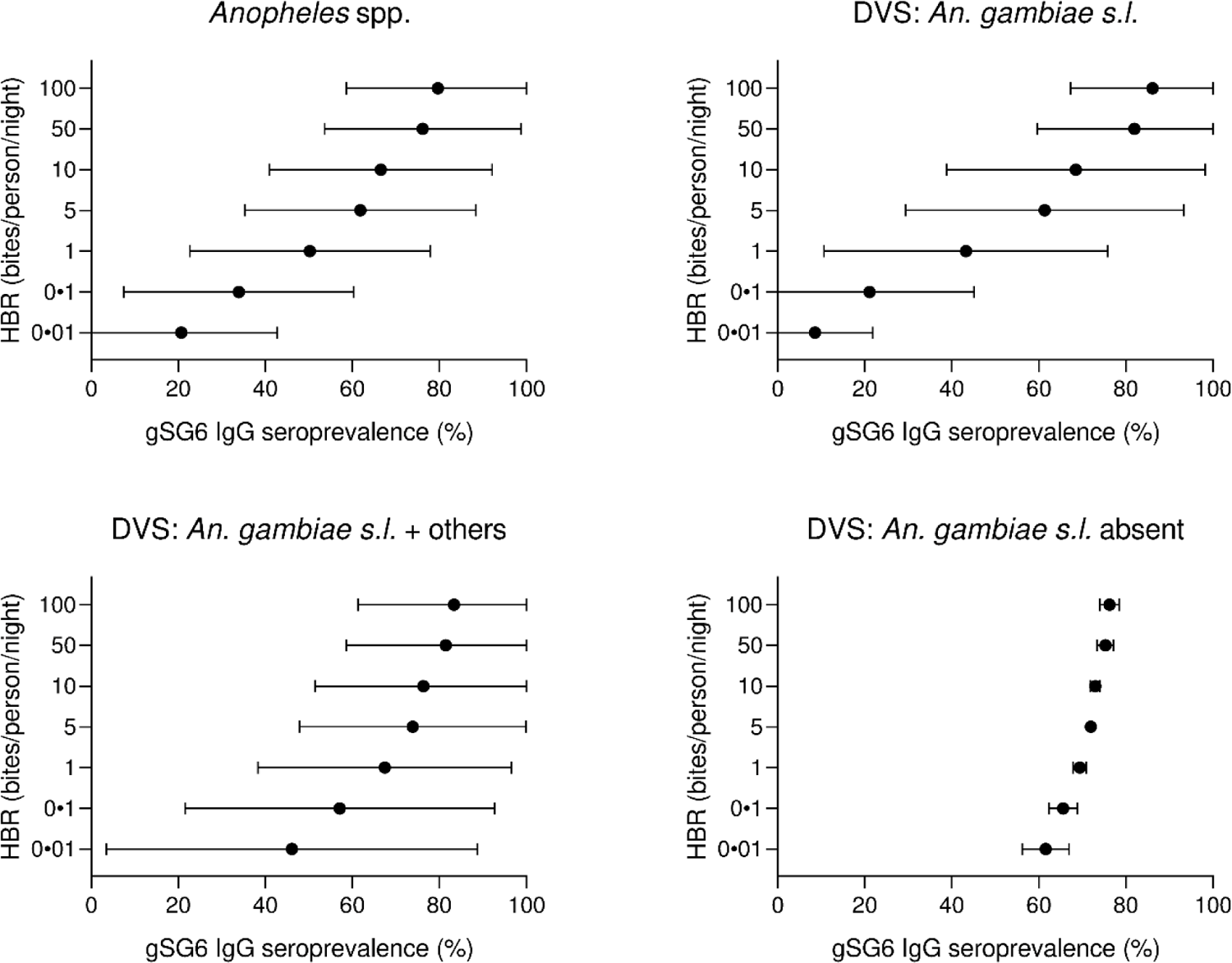
Forest plots of predicted anti-gSG6 IgG seroprevalence (%) and *Anopheles* species-specific human biting rate (HBR). Panels show the predicted average anti-gSG6 IgG seroprevalence (predicted probability for the average study and country) with 95% confidence intervals for given HBR, for all *Anopheles* spp. (using model output from Figure 2 – Supplement 1) and for specific-dominant vector species (DVS): where *An. gambiae s.l.* is the only DVS, where other DVS were present in addition to *An. gambiae s.l.* and where *An. gambiae s.l.* was absent (using model output from Figure 4 – Supplement 1).

A positive association was also found between seroprevalence of anti-gSG6 IgG antibodies and EIR in analysis of 38 observations from eight studies (Figure 3B, Figure 3 – Supplement 1) [3, 9, 13, 29, 39-41, 53]. For a 50% increase in EIR, the odds of anti-gSG6 IgG seropositivity increased by 6% (OR: 1.06; 95%CI: 1.03-1.10; *p*<0.001), with heterogeneity in the study-specific effects (95% reference range: 1.00-1.13; likelihood ratio χ^2^ (1) = 15.02, *p*<0.001). There was no evidence of effect modification by either vector collection method (*p*=0.095) or DVS (*p*=0.800) on the association between seroprevalence of anti-gSG6 IgG and EIR.

Similar positive associations were also found between anti-gSG6 IgG levels, HBR and EIR in 11 studies [7-11, 29, 36, 37, 39, 42, 52] and three studies [9, 13, 39] respectively but seven studies showed no association between HBR and levels of IgG to gSG6 [5, 13, 32, 33, 38, 43, 53].

The association between anti-gSG6 IgG seroprevalence and population-level prevalence of *Plasmodium* spp. infection was investigated. Generalised multilevel modelling of 212 meta-observations from 14 studies [3, 5, 9, 13, 15, 29, 30, 33, 35, 36, 38, 41, 47, 51] showed that for a 10% relative increase in the prevalence of *Plasmodium* spp. infection the odds of gSG6 IgG seropositivity increased by 4%, although the confidence intervals were wide (OR: 1.04; 95%CI: 0.98-1.11; *p*=0.148) (Figure 5 and Figure 5 – Supplement 1). In the association between gSG6 IgG seropositivity and *Plasmodium* spp. infection, there was no evidence for a moderating effect of *Plasmodium* spp. detection method (light microscopy, or PCR, *p*=0.968) or species (*P. falciparum* and *P. vivax,* or *P. falciparum*, *p*=0.586) potentially because all studies that reported *P. vivax* were co-prevalent with *P. falciparum*.

**Figure 5.**
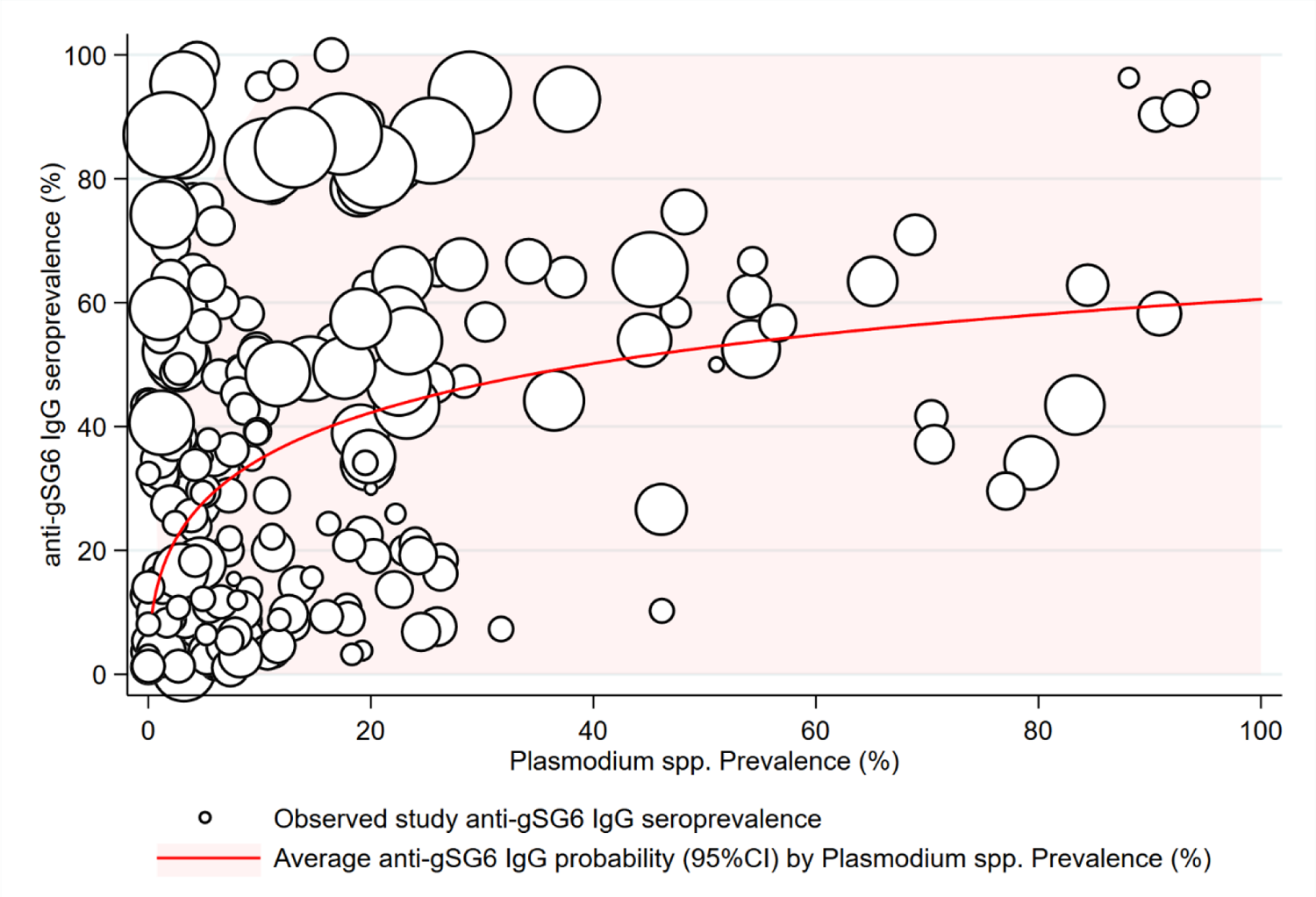
The association between anti-gSG6 IgG seroprevalence (%) and *Plasmodium* spp. prevalence (%). Figure shows the observed anti-gSG6 IgG (either recombinant or peptide form) seroprevalence (%) and prevalence of any *Plasmodium* spp. infection (%) for each meta-observation, as well as the predicted average anti-gSG6 IgG seroprevalence (predicted probability for average study) with 95% confidence intervals (95%CI). Circles are proportional to the size of the sample for each meta-observation. Association estimated using generalised multilevel mixed-effects modelling to account for the hierarchical nature of the data, using an anti-gSG6 IgG meta-observation, nested within study. See Figure 5 – Supplement 1 for model output.

Additionally, 14 studies reported estimates of anti-gSG6 IgG levels and the prevalence of *Plasmodium* spp. infections. The median anti-gSG6 IgG antibody levels increased with increasing *Plasmodium* spp. prevalence in six of these studies [5, 13, 15, 37, 38, 51], or in *Plasmodium* spp. infected compared to non-infected individuals [12, 50], but showed no association in eight studies [9, 29, 30, 32, 33, 35, 36, 43]. Furthermore, we also investigated associations with serological measures of malaria exposure and found that for a 10% relative increase in pre-erythrocytic and blood-stage stage antigen seroprevalence there was an 11% (OR: 1.11; 95%CI: 1.02-1.21%, *p*=0.013) and 5-27% (OR range: 1.05-1.27; *p* range: <0.001 to 0.523) increase in the odds of anti-gSG6 IgG seropositivity, respectively (Supplementary File 3).

To give epidemiological context we estimated anti-gSG6 seroprevalence by producing model-based predicted probabilities by malarial endemicity class. Multilevel modelling on 297 meta-observations from 22 studies shows that the estimated anti-gSG6 IgG seroprevalence is higher for the higher endemicity classes (eliminating malaria: 20% (95%CI: 8-31%); hypoendemic: 34% (95%CI: 19-49%); mesoendemic: 52% (95%CI: 35-68%); hyperendemic settings: 47% (95%CI: 27-64%); holoendemic: 78% (95%CI: 67-90%); *p*<0.001; Table 2). In addition, using Bayes Best-Linear-Unbiased Predictions (BLUPs) we estimated country-specific gSG6 IgG seroprevalence from an intercept only multilevel model fitted to 301 meta-observations from 22 studies. It showed that IgG seroprevalence to *An. gambiae* gSG6 was lowest in countries in the Pacific Region where *An. gambiae* is absent (Vanuatu (31%) and Solomon Islands (32%)) and highest in countries where *An. gambiae* is a DVS (Benin (72%) and Burkina Faso (65%); Supplementary File 4).

**Table 2:** Association between gSG6 IgG seroprevalence (%) and malarial endemicity (*Pf*PR_2-10_).

| Malaria Endemicity Class<sup>a</sup> | OR | 95%CI | $p$-value | Predicted gSG6 IgG seroprevalence (%) | 95%CI |
| --- | --- | --- | --- | --- | --- |
| *Eliminating malaria* ($PfPR <1\%$) | **Ref.** | | | 20.0 | 8.3, 31.7 |
| *Hypoendemic* ($PfPR 1-10\%$) | **2.04** | 1.43, 2.90 | $<0.001$ | 33.7 | 18.9, 48.5 |
| *Mesoendemic* ($PfPR 10-50\%$) | **4.19** | 2.80, 6.08 | $<0.001$ | 51.5 | 34.6, 67.7 |
| *Hyperendemic* ($PfPR 50-75\%$) | **3.36** | 1.98, 5.71 | $<0.001$ | 46.5 | 27.4, 63.8 |
| *Holoendemic* ($PfPR >75\%$) | **14.4** | 9.72, 21.36 | $<0.001$ | 78.2 | 66.8, 89.7 |

Table shows the odds ratio (OR), 95% confidence interval (95%CI), $p$ -value, as well as the predicted gSG6 IgG seroprevalence and associated 95%CI<sup>b</sup> for associations between endemicity class (derived from *P. falciparum* parasite rates in 2-10 year olds ( $PfPR_{2-10}$ )) and anti-gSG6 IgG seropositivity.
<sup>a</sup> Generalised multilevel modelling estimating the association between anti-gSG6 IgG seropositivity and endemicity class with random-effects for study-specific heterogeneity in gSG6 IgG. Model fitted to $n=297$ meta-observations. Endemicity class membership is derived from $PfPR_{2-10}$ from MAP, using cut-offs taken from Bhatt *et al.* [20], or where MAP data were unavailable, endemicity was included as indicated in the study.
<sup>b</sup> Predicted gSG6 IgG seroprevalence (predicted probability in the average study) is estimated from generalised multilevel modelling

Assessments of internal and external study validity revealed there was a moderate risk of selection bias (Supplementary File 2) due to the study-specific inclusion criteria of populations at higher risk of malaria which contributed gSG6 seroprevalence estimates.

## Discussion

This systematic review and multilevel modelling analysis provides the first quantification of a positive non-linear association between seroprevalence of *An. gambiae* gSG6 IgG antibodies and HBR and demonstrated that its magnitude varied with respect to the DVS present in the area. Importantly, this review identified a paucity of studies conducted outside of Africa, as well as investigating salivary antigens representing different *Anopheles* spp. and antigenic targets. gSG6 antibodies were positively associated with the prevalence of *Plasmodium* spp. infection as well as established epidemiological measures of malaria transmission: malaria endemicity class and EIR. Overall, our results demonstrate that antibody seroprevalence specific for *Anopheles* spp. salivary antigens has the potential to be an effective measure of vector exposure and malaria transmission at the population- and, potentially, individual-level.

*An. gambiae* gSG6 IgG seropositivity increased with increasing HBR, although these increases had diminishing impact on *An. gambiae* gSG6 IgG seropositivity at higher levels of HBR. We also observed that the association was strongest in areas where *An. gambiae s.l.* was the only DVS (that is concordant *An. gambiae* species-specific HBR with *An. gambiae* gSG6 antibodies). Associations, albeit weaker, were also observed between discordant species-specific HBR and gSG6, most likely because the *An. gambiae SG6* gene shares moderate sequence identity with vector species that are dominant in other regions (Africa: 80% *An. funestus*; Asia: 79% *An. stephensi* and *An. maculatus*; 54% *An. dirus*; Pacific: 52.5% *An. farauti*), and is absent from the DVS of the Americas (*An. albimanus* and *An. darlingi*) [55]. The generalisability of *An. gambiae* gSG6 IgG as a biomarker of exposure to other *Anopheles* spp. may therefore be limited. However, our review also identified a paucity of studies investigating additional salivary antigenic targets and *Anopheles* species not present in Africa. The identification of novel salivary antigens that are species-specific will be valuable in quantifying exposure to the other *Anopheles* vectors that share limited identity with *An. gambiae SG6* (such as *An. farauti* and *An. dirus*), as well as *Anopheles* spp. which lack *SG6* (as done for *An. albimanus* and *An. darlingi* [49, 56]). An *Anopheles* species-specific serological platform could advance vector surveillance by more accurately capturing exposure to DVS in the South American and Asia Pacific regions which exhibit diverse biting behaviours and vector competence (DVS typically bite outdoors during the night and day respectively [19, 57–61]), as well as the increasing threat of urban malaria from *An. stephensi* in Africa [62, 63].

Importantly, this review demonstrated that the prevalence of *Anopheles* salivary antibodies increased with increasing prevalence of *Plasmodium* spp. infection as well as established epidemiological measures of malaria transmission: malaria endemicity class and EIR. Anti-salivary antibodies, such as SG6 IgG, therefore, have the potential to serve as a proxy measure for vectorial capacity and malaria receptivity of a population to sustain malaria transmission. Their application could be particularly relevant in pre-elimination areas, or non-endemic areas under threat of imported malaria, where *Anopheles* salivary antibodies are more readily detectable than parasites; salivary antibodies were predicted to be prevalent (20%) in areas defined as eliminating malaria (<1% *Pf*PR_2-10_). Furthermore, if SG6 IgG seroprevalence can be effectively combined with a measurement of the sporozoite index, salivary antibodies as a marker of HBR could help overcome sensitivity limitations of EIR in low transmission areas. Additional measures could include estimates of malaria prevalence or serological biomarkers that are species- or life stage-specific (e.g. *Plasmodium* spp. pre-erythrocytic antigens as biomarkers for recent parasite inoculation). Indeed, positive associations between antibodies specific for *Plasmodium* spp. pre-erythrocytic and blood-stage antigens with gSG6 were demonstrated in analyses of data from diverse malaria endemic areas. Serological tools combining salivary antigens with antigens specific for the different *Plasmodium* spp. would be particularly beneficial in the Asia Pacific, a region of relatively low malaria transmission with goals of elimination, but the highest burden of *P. vivax* malaria where blood-stage infection can be caused by relapses from dormant liver stages. In these areas, parasite prevalence may therefore overestimate ongoing malaria transmission, making vector surveillance tools essential to informing elimination strategies in the Asia Pacific and other regions where *P. vivax* is endemic.

The gold standard entomological measures HBR and EIR provide crude population-level estimates of vector and malaria exposure that are specific in space and time and preclude investigation of individual-level heterogeneity and natural transmission dynamics. Our study demonstrated that salivary biomarkers measured at the individual-level, such as gSG6 IgG, can be used to quantify total vector exposure at the population-level, without requiring laborious entomological experiments. However, validating an individual-level serological measure, which demonstrates considerable individual-level variation, against the imperfect population-level gold standards of HBR and EIR is challenging and reflected in the variation in study-specific estimates in the association between gSG6 IgG and HBR in modelling analyses. However, the accuracy of salivary antibodies to measure individual-level exposure to *Anopheles* bites is yet to be validated; literature searches identified no studies investigating this association at the individual-level. Without detailed measurements of individual-level vector exposure, or a detailed knowledge of the half-life of *Anopheles* salivary antibodies post biting event, the true accuracy of salivary antibodies, such as SG6 IgG, to measure individual-level HBR remains unknown. This knowledge is particularly pertinent where *Anopheles* salivary biomarkers might be applied to assess the effectiveness of a vector control intervention or used to measure temporal changes in malaria transmission; particularly in areas or populations where there is considerable heterogeneity in individual-level risk of *Anopheles* exposure (*e.g.* unmeasured outdoor biting due to occupational exposure for forest workers [64]).

The broad nature of our inclusion and quality criteria was a key strength of our systematic review, which aimed to provide a comprehensive analysis of all *Anopheles* salivary biomarkers and determine their associations with entomological and malariometric measures of transmission. However, this review has two main limitations. First, despite the inclusive nature, assessment of the external validity of the review revealed a moderate risk of bias; some studies exhibited a high risk of selection bias as they were performed in specific high-risk populations not representative of the overall population (*i.e.* children only). This is accounted for to some degree by specification of a random effect (*i.e.* intercept) for study, which accounts for unmeasured study-specific factors that may introduce study-specific measurement error to measurement of the outcome. Second, with respect to internal validity, there may be potential selection bias introduced by the exclusion of studies reporting zero HBR (seven observations from three studies [9, 36, 53]), EIR (22 observations from three studies [9, 13, 29]) and malaria prevalence (15 observations from three studies [15, 36, 51]) estimates, given we modelled the log of these factors. However, adding a small constant (*e.g.* 0.001) to a zero value to permit modelling of a log estimate can also introduce considerable bias (*i.e.* seemingly small differences between values become very large on the log scale). In light of this, we also chose to provide estimates of association and gSG6 IgG seroprevalence according to widely accepted, discrete, endemicity classes (which permitted inclusion of all studies) and according to a selected range of epidemiologically relevant hypothetical HBR’s (no widely accepted HBR classification exists in the literature) to provide epidemiological context.

## Conclusions

In order to advance progress towards malaria elimination the World Health Organisation has called for innovative tools and improved approaches to enhance vector surveillance and monitoring and evaluation of interventions [65]. Our systematic review has provided strong evidence that *Anopheles* salivary antibodies are serological biomarkers of vector and malaria exposure, by quantifying their strong positive association with *Anopheles*-HBR and established epidemiological measures of malaria transmission. These salivary biomarkers have the potential to replace crude population-level entomological experiments with a precise and scalable tool that measures *Anopheles* vector exposure at the individual-level. This approach could be expanded into a sero-surveillance tool to assess the effectiveness of vector control interventions, define heterogeneity in malaria transmission and inform efficient resource-allocation, that would ultimately accelerate progress towards elimination.

## Contributors

All authors developed the protocol and the analytical plan. EAK performed literature searches, screened studies and extracted data with input from FJIF. EAK and PAA analysed the data. All authors interpreted the data. EAK and FJIF drafted the report. All authors read and critically revised the draft report, and approved the final report. All authors agreed to be accountable for all aspects of the work.

## Data Availability

All datasets and code associated with the analyses can be found at the https://github.com/ellenakearney/Anopheles_salivary_biomarker_systematic_review.git

## Acknowledgements

We wish to thank the authors of the original studies for responding to requests for further information/data for inclusion in the systematic review. This work was supported by the National Health and Medical Research Council of Australia (Australian Centre for Research Excellence in Malaria Elimination (ACREME) to FJIF and JAS (1134989); Career Development Fellowship to FJIF (1166753) and investigator award to JAS (1196068); and its Independent Research Institute Infrastructure Support Scheme), the Australian Commonwealth Government (Australian Government Research Training Program Scholarship awarded to EK) and a Victorian State Government Operational Infrastructure Support Program received by the Burnet Institute. This research was funded in part by the Wellcome Trust (220211). For the purpose of Open Access, the authors have applied a CC BY public copyright licence to any Author Accepted Manuscript version arising from this submission. The funders had no role in the study design, data collection or analysis, decision to publish, or preparation of the manuscript.

## Supplementary Files

Supplementary File 1. Supplementary Methodology.

### Search strategy

We performed a systematic review with multilevel modelling of the published literature according to the Meta-analysis of Observational Studies in Epidemiology (MOOSE) guidelines [1] and the Preferred Reporting Items for Systematic Reviews and Meta-Analyses (PRISMA) specifications [2]. The protocol was registered with PROSPERO (CRD42020185449).

The electronic databases PubMed, Scopus, Web of Science, African Index Medicus, and the Latin American and Caribbean Health Sciences Literature (LILACS) were searched for studies published before June 30, 2020 investigating *Anopheles* salivary antigens as a biomarker for mosquito exposure or malaria transmission. Search terms were as follows: *Anophel*\* AND saliva* AND (antibod* OR sero* OR antigen OR marker* OR biomarker* OR gSG6* OR gSG* OR SG* OR cE5). The reference lists of included studies were screened for additional studies, and Google Scholar was used to identify additional works by key authors. No formal attempt was made to identify unpublished population studies as it would have required significant description of the design, methods and analysis used in these studies, and a review of ethical issues.

### Selection criteria

The primary criteria for inclusion in this systematic review was the reporting of estimates of seroprevalence or total levels of Immunoglobulin (Ig) antibodies (including all isotypes and subclasses) in human sera against recombinant or synthetic peptide *Anopheles* salivary antigens. We considered for inclusion: cross-sectional studies, cohort studies, intervention studies and case-control studies of individuals or populations (including sub-populations) living in all geographies with natural exposure to *Anopheles* mosquitoes. Studies that were solely performed in participants not representative of the wider population (i.e. mosquito allergic patients, soldiers, returned travellers) were excluded. The minimum quality criteria for inclusion in this review were: antibody detection performed using enzyme-linked immunosorbent assay (ELISA), multiplex or Luminex assays.

The exposure variables of interest included entomological and malariometric parameters, including: (i) human biting rate (HBR), defined as the number of bites received per person per unit of time; (ii) entomological inoculation rate (EIR), defined as the number of infectious bites per person per unit of time, calculated as the HBR multiplied by the sporozoite index; (iii) estimates of malaria prevalence; (iv) population-level seroprevalence estimates against *Plasmodium* spp. malarial antigens. To ensure HBR estimates were given for the same unit of time (bites per person per night), biting rates given per week were divided by 7, and biting rates given per month we multiplied by 12 and divided by 365. Similar approaches were employed to ensure consistent units for EIR (infectious bites per person per year). *Plasmodium* spp. infections had to be confirmed by either microscopy, rapid diagnostic test (RDT) or molecular methods (polymerase chain reaction (PCR)). *Plasmodium* spp. diagnosis was included for all *Plasmodium* spp. combined and the species-level if provided. Where exposure estimates were not provided, we attempted to source data from other publications by the authors, or using the site geolocation and year to obtain estimates of EIR from the Pangaea dataset [3]. *P. falciparum* rates in 2-10 year olds (globally, 2000–2017) and dominant vector species (DVS) from the Malaria Atlas Project (MAP) [4]. Studies of salivary antigens where exposure variables could not be sourced and data that could not be extracted were excluded.

### Selection of studies

One author performed database searches and screened reference lists to identify possible studies. One author screened studies against inclusion criteria, with discussion and input from a second reviewer.

### Approaches to include all available studies

The authors of any studies that did not contain relevant information on the study design, populations, eligibility criteria, or key study data, were contacted and relevant data requested. Authors were contacted via an initial email detailing the precise nature of the systematic review and the data required. If the authors did not reply to three email requests, or were unable to provide relevant data, the studies were deemed to insufficiently meet inclusion/quality criteria and were excluded. As measurement of antibody levels does not produce a common metric between studies, authors were asked to classify their participants as ‘responders’ or ‘no-responders’ according to seropositivity (antibody level relative to unexposed sera) within each study, to allow comparisons of seroprevalence between studies [5–7]. Studies that were only able to provide antibody levels or categorised seropositivity based upon arbitrary cut offs were excluded from multilevel modelling analyses and included in narrative terms. Where the salivary antibody response and exposure variable were measured in the same population and reported in multiple publications, the study with the largest sample size was included, otherwise the earliest study was included.

### Data extraction

Data were extracted using a data collection form by one reviewer. Any data that was provided at the sub-population level was extracted at the lowest level i.e. if a study was performed across multiple sites, and an estimate for both salivary antibody seroprevalence/levels and the outcome of interest is given for each site, it was included the site level, rather than an aggregated level.

### Measures Outcomes

The primary outcome of interest of our systematic review was the reported antibody response (both seroprevalence and levels of all Ig subclasses and isotypes) to *Anopheles* salivary antigens. Multilevel modelling analyses were performed where the seroprevalence of antibodies against the same antigen and the exposure of interest were reported in more than one study.

### Exposures

The primary exposures of interest included in the multilevel modelling analyses were the HBR and EIR, a measure of the average number of bites received per person per night and infectious bites received per person per year, respectively. Secondary exposures assessed include the prevalence of any *Plasmodium* spp. infection (including

*P. falciparum* only, *P. vivax* only, or untyped infections). Additional secondary exposures include the *P. falciparum* infection rate in 2-10 year olds extracted from MAP, as well as the seroprevalence of antimalarial antibodies against pre-erythrocytic and blood-stage antigens.

Clinical and methodological heterogeneity were explored using prespecified variables to minimize spurious findings. Variables considered for inclusion were study design (cohort, cross-sectional, repeated cross-sectional), DVS, study participants (adults only, children only, adults and children), preparation of salivary antigen (recombinant full-length protein, synthetic peptide), malaria detection methodology (light microscopy, RDT, PCR), and entomological vector collection methodology (human landing catch, light traps, and spray catches).

### Statistical analysis

Where there were sufficient data to pool estimates of the same exposure and outcome measures, generalised linear mixed modelling (GLMM) was used to undertake analyses quantifying associations between the exposures of interest and salivary antibody seroprevalence measurements. Models were generalised through use of the logit link function and binomial distribution (statistical notation for HBR model shown below as equation one). Seroprevalence was modelled in binomial form as the number of individuals seropositive to the total sample size. A three-level random effects model with a nested framework was used to account for dependency in the data, with random intercepts for country (level-3) and study (level-2) and level-1 units representing multiple observations per study induced by the study design (*i.e.* multiple time points, sites, age categories). Additionally, random slopes for entomological and malariometric exposure parameters were included to permit the effect of the exposure of interest (HBR/EIR/malaria prevalence/etc.) to vary across studies. Model structure was determined empirically through likelihood ratio tests (*p*<0.05), with the exception of country at the 3rd level which was included in HBR and EIR analyses to estimate country-specific seroprevalence estimates of anti-salivary antibodies. The associations between the various exposures and the different salivary antigens were analysed separately, however estimates of IgG seroprevalence against the recombinant full-length protein (gSG6) and synthetic peptide (gSG6-P1, the one peptide determined in all studies utilising peptides) form of the gSG6 antigen were analysed together, with a fixed term for antigen construct considered for inclusion in the model. Of note, gSG6 peptide 2 (gSG6-P2) was excluded from being analysed with gSG6 and gSG6-P1 estimates, as the two studies that reported anti-gSG6-P2 IgG seroprevalence also reported the seroprevalence of anti-gSG6-P1 IgG, and only one could be included. Potential effect modification of the associations between the exposures of interest and the anti-*Anopheles* salivary antibody responses was explored was undertaken by estimating interaction terms for DVS (*An. gambiae sensu lato* (*s.l.*) only, *An. gambiae s.l.* and other DVS, or *An. gambiae s.l.* absent) and for vector collection method (human landing catch or other indirect measures *e.g.* light traps, spray catches, etc.). For the association between *Plasmodium* spp. prevalence and gSG6 IgG seropositivity interaction terms for malaria detection methodology (light microscopy or PCR), and malarial species type (*P. falciparum* only, or *P. falciparum* and *P. vivax*) were estimated. Other variables considered for inclusion in adjusted models were study design, participant, salivary antigen construct; however, these variables showed no association with anti-gSG6 IgG and were thus excluded.

The natural log of HBR, EIR, malaria prevalence and antimalarial antibody seroprevalence were modelled to account for the non-linear functional form of the association between each measure and the log odds of anti-gSG6 IgG seroprevalence - supported empirically by superior model fit as indicated by Akaike’s information criterion (AIC) and Bayesian information criterion (BIC) fit indices. To aid interpretation, odds ratios are presented according to relative per cent increase in the exposures, such that odds ratios reflect relative changes in anti-gSG6 IgG seropositivity for 50% increases HBR and EIR, and 10% increases in malaria prevalence and antimalarial seroprevalence. Additional per cent changes in HBR and EIR are also presented.

Empirical Bayes best linear unbiased predictions (BLUPs) were used to estimate the probability of gSG6 IgG seropositivity in the average study and country, which is equivalent to an estimated gSG6 IgG seroprevalence. In order to maximise the number of included studies in our modelling, we predicted anti-gSG6 seroprevalence according to endemicity class, derived by applying established endemicity cut-offs to *Pf*PR_2-10_ estimates [8] extracted from MAP using site year and geolocation (if MAP data unavailable endemicity as stated in study). Intraclass correlation coefficients (ICCs) and 95% reference ranges were estimated for country-, study- and slope-specific heterogeneity (where appropriate) using estimated model variance components.

*Statistical notation for the generalised linear mixed model used to estimate the association between Anopheles gambiae gSG6 IgG seropositivity and human biting rate (HBR)*.

The model can be formally written as:

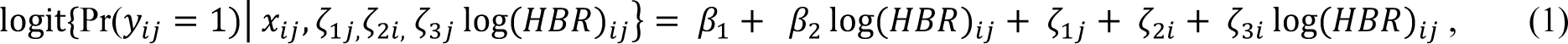

where

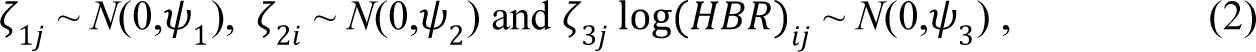

Where x_ij_ is the vector of model covariates, β_1_ is the model constant and represents the log odds (probability) of gSG6 IgG seropositivity for a log HBR of zero, β_2_ the fixed effect for log HBR for country *j* and study *i*, ζ_1j_ the random-effect (i.e. intercept) for between-country heterogeneity in probability of gSG6 IgG seropositivity, ζ_2i,_ the random-effect (i.e. intercept) for between-study heterogeneity in probability of gSG6 IgG seropositivity, and ζ_3i_ the random-effect (i.e. coefficient) for between-study heterogeneity in the effect of log HBR.

### Risk of bias in individual studies

For cross-sectional, cohort or intervention studies, selection bias was assessed by reviewing the studies’ inclusion and exclusion criteria. Any case-control studies, or studies that presented salivary antibody data stratified by malaria infection status were included in narrative terms only. Risk of bias was assessed by one reviewer using the Risk of Bias in Prevalence Studies tool [9]. The risk of bias pertains to the reported seroprevalence estimates of anti-*Anopheles* salivary antibodies included in the multilevel modelling.

### Supplementary File 2. Risk of Bias assessment

Risk of bias was assessed for each study by one independent reviewer using the *Risk of Bias in Prevalence Studies* tool [9]. This tool comprises 10 items and a summary assessment to assess the external validity (selection and non-response bias) and internal validity (measurement bias) of the study’s prevalence estimates. The risk of bias pertains to the reported seroprevalence estimates of anti-*Anopheles* salivary antibodies included in the multilevel modelling.

With regard to external validity, seven of the studies included in the review were performed in specific populations (*i.e.* children only) that were not representative of the national population and were deemed to be at high risk of selection bias. Only 7 studies included some form of random sampling, and frequently insufficient detail was provided on the sampling frame; as such most studies were included as high risk of selection bias. Furthermore, no studies reported participant response-rate, and as such were indicated as high risk of nonresponse bias.

In terms of internal validity, all studies had an acceptable case definition, with the same mode of data collection, a valid instrument and an acceptable prevalence period, so were all deemed to be of low risk. However, 12 studies did not include a denominator, instead only reporting the study sample size and prevalence estimate, and were included as high risk.

Overall, due to the specific nature of some of the sample populations for which these prevalence estimates are given (*i.e.* children only) and as participant non-response rate is not given, we conclude that there is a moderate risk of study bias. According to the *Risk of Bias in Prevalence Studies* tool [9], this implies that future research is likely to have an impact on our confidence in the prevalence estimates.

### Supplementary File 3. Association between gSG6 IgG seropositivity and antimalarial antibody seroprevalence

#### Antibodies against *P. falciparum* pre-erythrocytic stage antigens

The pooled analysis of 159 meta-observations from eight studies showed that a 10% relative increase in PfCSP IgG seropositivity was associated with a 11% (OR: 1.11; 95%CI: 1.02-1.21%, *p*=0.013) increase in odds of anti-gSG6 IgG seropositivity [126–133]. Furthermore we observed that gSG6 IgG levels increased with increasing PfCSP IgG seroprevalence in four studies [127–129, 133], with another study contributing only a single estimate [132].

#### Antibodies against *P. falciparum* blood stage antigens

Furthermore, we observed a 10% relative increase PfAMA1 IgG seroprevalence was associated with a 13% (OR: 1.13; 95%CI: 1.12-1.15%; *p<*0.001) increase in odds of gSG6 IgG seropositivity, based upon 62 meta-observations from eight studies [128–135]. A similar association was observed for PfMSP1_19_ IgG, with 10% relative increase in seroprevalence associated with 13% (OR: 1.13; 95%CI: 1.03-1.25%; *p*=0.014) increase in odds of gSG6 IgG seropositivity. This association was derived from 163 meta-observations from ten studies [127-130, 132-136]. Analysis of 47 meta-observations from three studies indicated that a 10% relative increase in PfMSP2 IgG seroprevalence was associated with a 5% (OR: 1.05; 95%CI: 1.03-1.07%; *p*<0.001) increase in odds of gSG6 IgG seropositivity [130, 133, 135]. While 17 meta-observations from two studies showed a 10% relative increase in PfMSP3 IgG seroprevalence was associated with a 14% (OR: 1.14; 95%CI: 1.13-1.16%; *p*<0.001) increase in odds of gSG6 IgG seropositivity [132, 135].

The pooled analysis of 128 meta-observations from five studies showed that a 10% relative increase in PfGLURP IgG seroprevalence was associated with a 17% (OR: 1.17; 95%CI: 1.14-1.19%; *p*<0.001) increase in odds of gSG6 IgG seropositivity [126–130]. And 18 meta-observations from five studies indicated that 10% relative increase in *P. falciparum* schizont extract IgG seropositivity was associated with a 27% (OR: 1.27; 95%CI: 0.61-2.65%; *p*=0.523) increase in odds of gSG6 IgG seropositivity [128-130, 134, 137].

We observed that increasing seroprevalence of IgG antibodies against PfAMA1 saw increased levels of anti-gSG6 IgG in three studies [128, 129, 134], but no association in another [133]. The levels of gSG6 IgG increased with increasing PfMSP1_19_ IgG seroprevalence in three studies [128, 134, 136], but showed no association in three other studies [127, 129, 133]. No association between gSG6 IgG levels and MSP2 IgG seroprevalence was observed in one study [133]. PfGLURP IgG seroprevalence and gSG6 IgG antibody levels were reported in three studies, with one study reporting increased levels [128], one study reporting no association [127], and one study reporting decreased levels of anti-gSG6 IgG with increasing anti-PfGLURP seroprevalence [129]. One study showed increasing gSG6 IgG levels with increasing *P. falciparum* schizont extract IgG, while three other studies showed no association [128, 129, 137]. Of note, one study provided a single seroprevalence estimate of antibodies against PfAMA1, PfMSP1_19_ and PfMSP3 so no relationships can be drawn [132].

### Antibodies against *P. vivax* antigens

In pooled analyses of 115 meta-observations from two studies [127, 134], we observed that 10% relative increase in the seroprevalence of PvAMA1 was associated with a 20% (OR: 1.20; 95%CI: 1.19-1.22%; *p*<0.001) increase in the odds of anti-gSG6 IgG seropositivity. Furthermore, in 103 meta-observations from two studies [127, 134], 10% relative increase in PvMSP1_19_ IgG seroprevalence was associated with a 13% (OR: 1.13; 95%CI: 1.12-1.13%; *p*<0.001) increase in the odds of anti-gSG6 IgG seropositivity. However, neither study showed an association between the levels of gSG6 IgG and the seroprevalence of PvAMA1 and PvMSP1_19_ IgG [127, 134].

### Supplementary File 4. Country and study-specific predicted probability of gSG6 IgG seropositivity

In order to obtain estimates of gSG6 IgG seroprevalence for each country and study, an intercept only three-level random effects logistic regression was fitted to 301 meta-observations from 22 studies. The predicted probability of gSG6 IgG seropositivity was calculated at the country-level (Figure S1), indicating that the seroprevalence was lowest in the Pacific Region (Vanuatu (31%) and Solomon Islands (32%)) and highest in Benin (72%) and Burkina Faso (65%). Furthermore, the predicted probability of gSG6 IgG seropositivity was calculated at the study-level (Figure S2) indicating that the seroprevalence was lowest in Ambrosino *et al.* [126] (13%) and highest in Drame *et al.* [138] (91%).

### Supplementary File 5. Association between alternative salivary biomarkers and exposures of interest

Our systematic review identified a paucity of studies that assessed the relationship between our exposures of interest and most alternate *Anopheles* salivary biomarkers (that is non-*An. gambiae* gSG6 IgG), thus preventing the estimation of a pooled association. The exceptions being that we observed that a 50% relative increase in HBR was associated with a 7% increase (OR: 1.07; 95%CI: 1.01-1.13%; *p*=0.017) in odds of anti-*An. funestus* fSG6 IgG seropositivity (six meta-observations from two studies [139, 140]; Table 1), as well as a 42% (OR: 1.42; 95%CI: 1.39-1.46%; *p*<0.001) and 21% (OR: 1.21; 95%CI: 1.19-1.23%; *p*<0.001) increase in odds of anti-gSG6-P2 IgG seropositivity associated with 10% relative increase in seroprevalence of PfCSP and PfGLURP IgG, respectively (115 and 116 meta-observations from two studies respectively [126, 127], Table 2-3). The associations between exposures of interest and the additional salivary biomarkers are further discussed in narrative terms in below.

#### Human biting rate

In addition to the increased odds of *An. funestus* fSG6 seropositivity with increasing HBR, the majority of studies reported a positive association between HBR and the seroprevalence and levels of anti-gSG6-P1 IgM [138], the levels of gSG6-P2 IgG [141], the seroprevalence and levels of anti-cE5 IgG [142], the levels of anti-fSG6 IgG [139, 140], the seroprevalence and levels of anti-f5’nuc IgG [139] and the median levels of anti-*An. gambiae* salivary gland extracts (SGE) SGE IgG and IgG4 [143–145]. One study reported similar median levels of anti-gSG6 IgG1 across populations and time points, whilst reporting that anti-gSG6 IgG4 titre increased with increasing HBR in one of the populations, but not in the other [146]. Similarly, there was no consistent association between HBR and the levels of anti-cE5 IgG [147], levels of anti-*An. gambiae* SGE IgE [145] and the seroprevalence and levels of anti-g5’nuc IgG [139].

### Entomological inoculation rate

Ali *et al.* [139] reported higher seroprevalence and levels anti-fSG6 IgG and anti-f5’nuc IgG with increasing EIR, while anti-g5’nuc IgG seroprevalence and levels were not associated with EIR. An additional study reported gSG6-P2 IgG seroprevalence estimates of 0% for three sites, irrespective of EIR [126].

### Malaria prevalence

Two studies showed that increased *Plasmodium* spp. prevalence was associated with higher median levels of anti-*An. gambiae* SGE IgG [143, 148], while another study showed different anti-*An. gambiae* SGE IgG levels for very similar prevalence of malaria and slightly lower levels of anti-*An. gambiae* SGE IgE and IgG4 for the time point with greater malaria prevalence [145]. Kerkhof *et al.* [127] showed increasing levels of anti-gSG6-P2 IgG for higher prevalence of any *Plasmodium* spp. infection, while Londono-Renteria *et al.* [149] showed lower levels of IgG antibodies against TRANS-P1, TRANS-P2, PEROX-P1, PEROX-P2 and PEROX-P3 in the site with higher PCR confirmed malaria prevalence. Additionally, several case-controlled studies, and two cross-sectional study, reported median antibody levels stratified by malaria infection status. These studies show higher levels of anti-*An. darlingi* SGE IgG [150], anti-*An. gambiae* SGE IgG [144], anti-*An. dirus* SGE IgG and IgM [151], and IgG antibodies against SGEs of two Colombian strains of *An. albimanus* in *Plasmodium* spp. infected individuals, compared to non-infected [152]. While Montiel *et al.* [152] observed no association between anti-*An. darlingi* SGE IgG levels and infection status.

### Antimalarial antibody seroprevalence

Our multilevel modelling indicated that there were 42% (OR: 1.42; 95%CI: 1.39-1.46%; *p*<0.001) and 21% (OR: 1.21; 95%CI: 1.19-1.23%; *p*<0.001) increase in odds of anti-gSG6-P2 IgG seropositivity associated with a 10% relative increase in the seroprevalence of PfCSP and PfGLURP IgG, respectively [126, 127]. However, we observed weak positive associations between the levels of IgG antibodies against gSG6-P2 peptide and the seroprevalence of IgG antibodies against PfMSP1_19_, PfGLURP and PvMSP1_19_, but no association with PfCSP or PvAMA1 [127].

**Figure S1:**
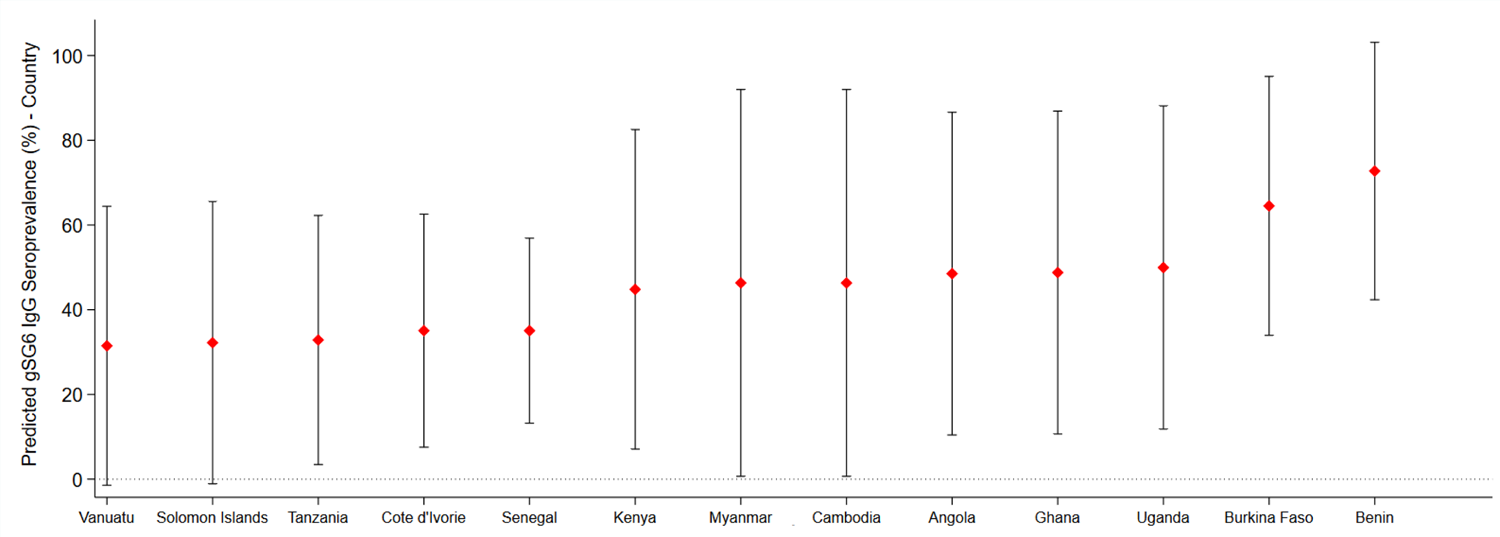
Risk of Bias assessment. Red – high risk, orange – moderate risk, green – low risk. Figure 1 - Supplement 1. Reasons for study exclusion.

**Figure S1:**
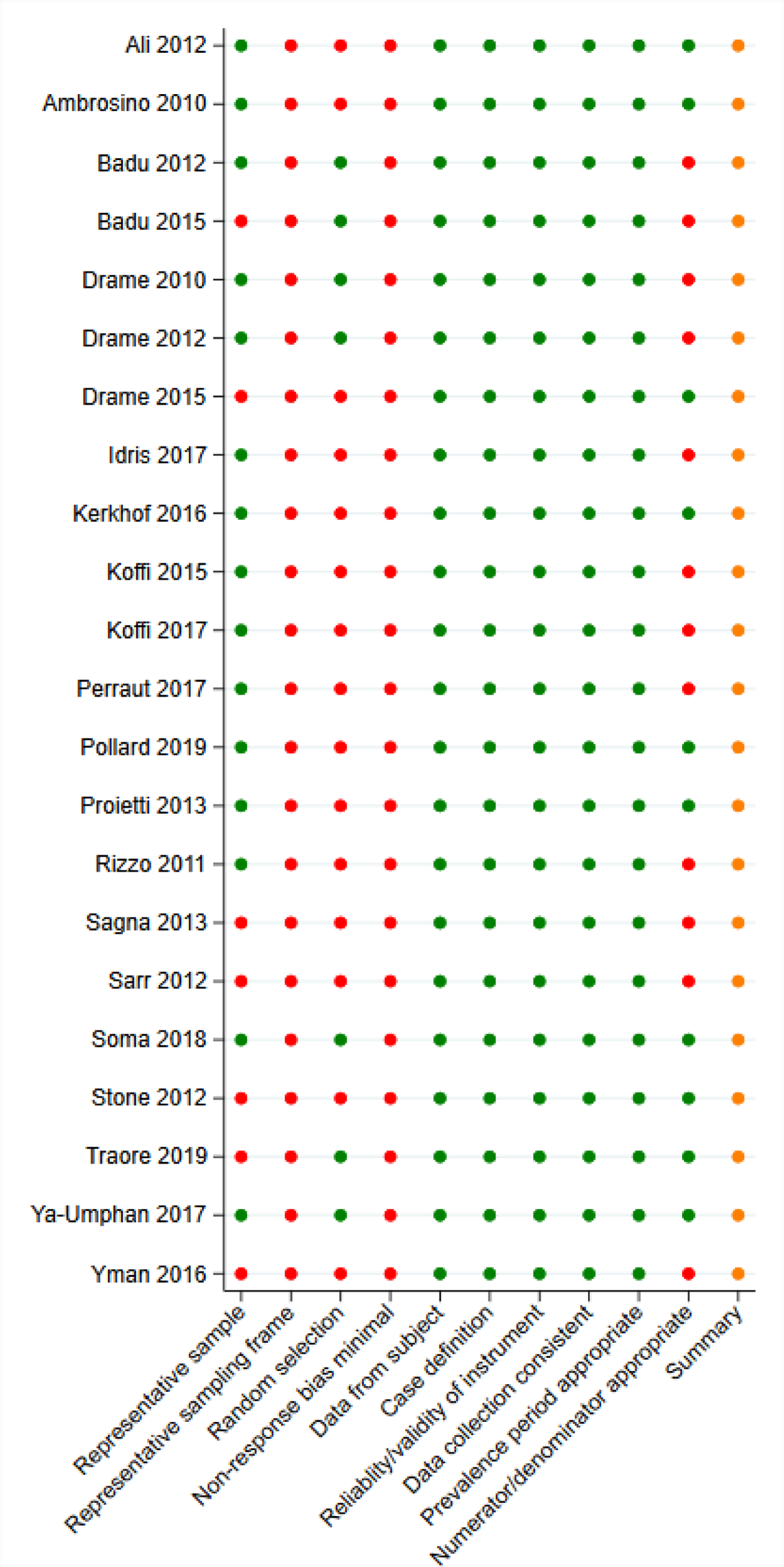
Predicted gSG6 IgG seroprevalence by country. Predicted probabilities of gSG6 IgG seropositivity including country-specific random effects with 95% confidence intervals. Estimated from intercept-only three-level random-effects logistic regression to account for the hierarchical nature of the data, with an anti-gSG6 IgG meta-observation nested within study nested within country. Based upon 301 meta-observations from 22 studies.

**Figure S2:**
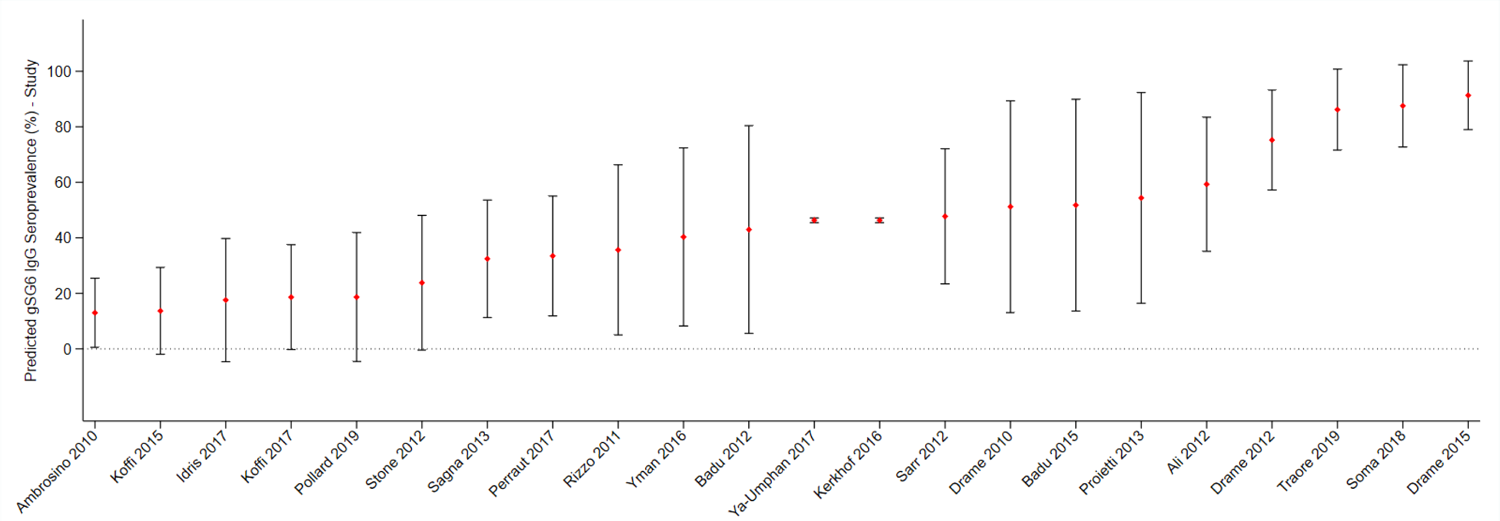
Predicted gSG6 IgG seroprevalence by study. Predicted probabilities of gSG6 IgG seropositivity including study-specific random effects with 95% confidence intervals. Estimated from intercept-only three-level random-effects logistic regression to account for the hierarchical nature of the data, with an anti-gSG6 IgG meta-observation nested within study nested within country. Based upon 301 meta-observations from 22 studies.

**Table S1:** Reasons for study exclusion Figure 2 **– Supplement 1: Association between gSG6 IgG seropositivity and human biting rate**

| Studies | Reason | References |
| --- | --- | --- |
| 30 | Does not measure anti-salivary antibody responses in individuals/populations | [10-39] |
| 28 | Review article | [40-67] |
| 20 | *Anopheles* salivary antigens not assessed | [68-87] |
| 10 | Wrong antibody detection methodologies | [88-97] |
| 7 | Grey literature | [98-104] |
| 6 | Not performed in humans | [105-110] |
| 4 | Data already captured by our review from another publication | [111-114] |
| 3 | Unable to determine appropriate exposure estimate | [115-117] |
| 3 | Not in population with natural exposure | [118-120] |
| 1 | Hypothesis study | [121] |
| 1 | Pooled sera | [122] |
| 1 | Does not provide estimate of seroprevalence/total levels of antibodies against salivary proteins | [123] |
| 1 | Study population not representative: Mosquito allergic patients | [124] |
| 1 | Study population not representative: Soldiers with transient exposure | [125] |

**Table S1:** **Unadjusted association between gSG6 IgG seropositivity and log Human Biting Rate (HBR).** Figure 3 – Supplement 1: Association between gSG6 IgG seropositivity and entomological inoculation rate

| *Variable* | *Log Odds Ratio (SE)* | *95% CI* | *p-value* | *RE* |
| --- | --- | --- | --- | --- |
| ***Fixed part*** |  |  |  |  |
| log HBR | 0.29 (0.08) | 0.14, 0.45 | <0.001 |  |
| ***Random part*** |  |  |  |  |
| $\psi_1^c$ | | | | 1.29 |
| $\psi_2$ | | | | 1.55 |
| $\psi_3$ | | | | 0.06 |
| $\rho_1^d$ | | | | 0.21 |
| $\rho_2^e$ | | | | 0.47 |
| $\ell$ | | | | -1489.9 |
| ***Model fit indices*** |  |  |  |  |
| AIC |  |  |  | 2989.9 |
| BIC |  |  |  | 3004.2 |

Human biting rate (HBR) association: log odds ratio and standard error (SE), 95% confidence interval (95%CI), p-value, random-effect components (RE): variances ( $\psi$ ), conditional intraclass correlation coefficient ( $\rho$ )<sup>a</sup> and model log likelihood ( $\ell$ ) from generalised multilevel modelling.<sup>b</sup> This analysis is based upon n=132 meta-observations.
<sup>a</sup> $\rho = \frac{\psi_k + \dots + \psi_{nk}}{\psi_k + \dots + \psi_{nk} + \pi^2/3}$ , where $\psi_k$ through $\psi_{nk}$ are random-effect variance estimates pertaining to each of the respective variance components (see table notes <sup>c-e</sup>) from the generalised multilevel modelling for a specific ICC estimate.
<sup>b</sup> Generalised multilevel modelling estimating the association between log transformed HBR and anti-gSG6 IgG seropositivity with random-effects for country-specific and study-specific heterogeneity in gSG6 IgG seroprevalence and study-specific heterogeneity in effect of HBR.
<sup>c</sup> $\psi_1$ , $\psi_2$ and $\psi_3$ represent variances of the random-effects for country, study and effect of HBR respectively.
<sup>d</sup> $\rho_1$ represents conditional ICC for meta-observations from the same country but different study.
<sup>e</sup> $\rho_2$ represents conditional ICC for meta-observations from the same country and study with the median HBR

**Table S1:** Unadjusted association between gSG6 IgG seropositivity log Entomological Inoculation Rate (EIR). Figure 4 – Supplement 1: Association between gSG6 IgG seropositivity and Human Biting Rate (HBR), moderated by dominant vector species

| *Variable* | *Log Odds Ratio (SE)* | *95% CI* | *p-value* | *RE* |
| --- | --- | --- | --- | --- |
| ***Fixed part*** |  |  |  |  |
| log EIR | 0.15 (0.04) | 0.07, 0.23 | <0.001 |  |
| ***Random part*** |  |  |  |  |
| $\psi_1^c$ | | | | 1.02 |
| $\psi_2$ | | | | 2.15 |
| $\psi_3$ | | | | 0.01 |
| $\rho_1^d$ | | | | 0.16 |
| $\rho_2^e$ | | | | 0.49 |
| $\ell$ | | | | -530.7 |
| ***Model fit indices*** |  |  |  |  |
| AIC |  |  |  | 1071.3 |
| BIC |  |  |  | 1079.5 |

Entomological inoculation rate (EIR) association: log odds ratio and standard error (SE), 95% confidence interval (95% CI), p-value, random-effect components (RE): variances ( $\psi$ ), conditional intraclass correlation coefficient ( $\rho$ )<sup>a</sup> and model log likelihood ( $\ell$ ) from generalised multilevel modelling.<sup>b</sup> This analysis is based upon n=38 meta-observations.
<sup>a</sup> $\rho = \frac{\psi_k + \dots + \psi_{nk}}{\psi_k + \dots + \psi_{nk} + \pi^2/3}$ , where $\psi_k$ through $\psi_{nk}$ are random-effect variance estimates pertaining to each of the respective variance components (see table notes <sup>c-e</sup>) from the generalised multilevel modelling for a specific ICC estimate.
<sup>b</sup> Generalised multilevel modelling estimating the association between log transformed EIR and anti-gSG6 IgG seropositivity with random-effects for country-specific and study-specific heterogeneity in gSG6 IgG seroprevalence and study-specific heterogeneity in effect of EIR.
<sup>c</sup> $\psi_1$ , $\psi_2$ and $\psi_3$ represent variances of the random-effects for country, study and effect of EIR respectively.
<sup>d</sup> $\rho_1$ represents the conditional ICC for meta-observations from the same country but different study.
<sup>e</sup> $\rho_2$ represents the conditional ICC for meta-observations from the same country and study with the median EIR

**Table S1:** Association between gSG6 IgG seropositivity and log Human Biting Rate (HBR), moderated by dominant vector species Figure 5 – Supplement 1. Association between gSG6 IgG seropositivity and malaria prevalence

| *Variable* | *Log Odds Ratio (SE)* | *95% CI* | *p-value* | *RE* |
| --- | --- | --- | --- | --- |
| **Fixed part** |  |  |  |  |
| log HBR | 0.46 (0.11) | 0.25, 0.66 | <0.001 |  |
| DVS |  |  | <0.001* |  |
| *An. gambiae s.l.* only | Ref. |  |  |  |
| *An. gambiae s.l.* & other DVS | 1.00 (0.18) | 0.65, 1.25 | <0.001 |  |
| Non-*An. gambiae s.l.* | 1.09 (0.68) | -0.24, 2.42 | 0.109 |  |
| log HBR by DVS |  |  | <0.001* |  |
| *An. gambiae s.l.* only | Ref. |  |  |  |
| *An. gambiae s.l.* & other DVS | -0.26 (0.08) | -0.41, -0.11 | 0.001 |  |
| Non-*An. gambiae s.l.* | -0.38 (0.11) | -0.59, -0.17 | <0.001 |  |
| **Random part** |  |  |  |  |
| $\psi_1^c$ | | | | 0.96 |
| $\psi_2$ | | | | 2.32 |
| $\psi_3$ | | | | 0.08 |
| $\rho_1^d$ | | | | 0.14 |
| $\rho_2^e$ | | | | 0.51 |
| $\ell$ | | | | -1488.8 |
| **Model fit indices** |  |  |  |  |
| AIC |  |  |  | 2995.53 |
| BIC |  |  |  | 3021.48 |

Human biting rate (HBR) X dominant vector species (DVS) association: log odds ratio and standard error (SE), 95% confidence interval (95%CI), p-value, random-effect components (RE): variances ( $\psi$ ), conditional intraclass correlation coefficient ( $\rho$ )<sup>a</sup> and model log likelihood ( $\ell$ ) from generalised multilevel modelling.<sup>b</sup> \*indicates p-value from joint Wald test for polytomous variables. This analysis is based upon n=132 meta-observations.
<sup>a</sup> $\rho = \frac{\psi_k + \dots + \psi_{nk}}{\psi_k + \dots + \psi_{nk} + \pi^2/3}$ , where $\psi_k$ through $\psi_{nk}$ are random-effect variance estimates pertaining to each of the respective variance components (see table notes <sup>c-e</sup>) from the generalised multilevel modelling for a specific ICC estimate.
<sup>b</sup> Generalised multilevel modelling estimating the association between log transformed HBR and anti-gSG6 IgG seropositivity including an interaction term between DVS and log HBR with random-effects for country-specific and study-specific heterogeneity in gSG6 IgG seroprevalence and study-specific heterogeneity in effect of HBR.
<sup>c</sup> $\psi_1$ , $\psi_2$ and $\psi_3$ represent variances of the random-effects for country, study and effect of HBR respectively.
<sup>d</sup> $\rho_1$ represents the conditional ICC for meta-observations from the same country but different study.
<sup>e</sup> $\rho_2$ represents the conditional ICC for meta-observations from the same country and study with the median HBR

**Table S1:** Unadjusted association between gSG6 IgG seropositivity and log *Plasmodium* spp. prevalence.

| *Variable* | *Log Odds Ratio (SE)* | *95% CI* | *p-value* | *RE* |
| --- | --- | --- | --- | --- |
| ***Fixed part*** |  |  |  |  |
| log *Plasmodium* spp. prevalence | 0.46 (0.32) | -0.16, 1.08 | 0.148 |  |
| ***Random part*** |  |  |  |  |
| $\psi_1^c$ | | | | 17.21 |
| $\psi_2$ | | | | 1.25 |
| $\rho_1^d$ | | | | 0.85 |
| $\ell$ | | | | -2597.2 |
| ***Model fit indices*** |  |  |  |  |
| AIC |  |  |  | 5202.47 |
| BIC |  |  |  | 5215.90 |

Any *Plasmodium* species infections (including prevalence estimates of *P. falciparum* only, *P. vivax* only, both *P. falciparum* and *P. vivax* and un-typed infections): log odds ratio and standard error (SE), 95% confidence interval (95%CI), p-value, random-effect components (RE): variances ( $\psi$ ), conditional intraclass correlation coefficient ( $\rho$ )<sup>a</sup> and model log likelihood ( $\ell$ ) from generalised multilevel modelling.<sup>b</sup> This analysis is based upon n=212 meta-observations.
<sup>a</sup> $\rho = \frac{\psi_k + \dots + \psi_{nk}}{\psi_k + \dots + \psi_{nk} + \pi^2/3}$ , where $\psi_k$ through $\psi_{nk}$ are random-effect variance estimates pertaining to each of the respective variance components (see table notes <sup>c-d</sup>) from the generalised multilevel modelling for a specific ICC estimate.
<sup>b</sup> Generalised multilevel modelling estimating the association between the log prevalence of any *Plasmodium* spp. infection and anti-gSG6 IgG seropositivity with random-effects for study-specific heterogeneity in gSG6 IgG seroprevalence and study-specific heterogeneity in effect of *Plasmodium* spp. prevalence.
<sup>c</sup> $\psi_1$ and $\psi_2$ represent variances of the random-effects for study and effect of *Plasmodium* spp. prevalence respectively.
<sup>d</sup> $\rho_1$ represents the conditional ICC for meta-observations from the same study and with the median *Plasmodium* spp. prevalence.

**Table S1:** Associations between anti-gSG6 IgG seropositivity and log of antimalarial antibody seroprevalence.

| *Exposure* | *Log Odds Ratio (SE)* | *95%CI* | *p- value* | *Meta-N* | *Studies* | *References* |
| --- | --- | --- | --- | --- | --- | --- |
| **Pre-erythrocytic antigens** |  |  |  |  |  |  |
| log PfCSP IgG Seroprevalence (%) | 1.13 (0.45) | 0.24, 2.01 | 0.013 | 159 | 8 | [126-133] |
| **Blood stage antigens** |  |  |  |  |  |  |
| log PfAMA1 IgG Seroprevalence (%)<sup>a</sup> | 1.30 (0.07) | 1.17, 1.44 | <0.001 | 62 | 8 | [128-135] |
| log PfMSP1<sub>19</sub> IgG Seroprevalence (%) | 1.31 (0.53) | 0.27, 2.36 | 0.014 | 163 | 10 | [127-130, 132-136] |
| log PfMSP2 IgG Seroprevalence (%) | 0.50 (0.11) | 0.27, 0.72 | <0.001 | 47 | 3 | [130, 133, 135] |
| log PfMSP3 IgG Seroprevalence (%)<sup>a</sup> | 1.41 (0.09) | 1.24, 1.58 | <0.001 | 17 | 2 | [132, 135] |
| log PfGLURP IgG Seroprevalence (%) | 1.61 (0.12) | 1.37, 1.85 | <0.001 | 128 | 5 | [126-130] |
| log PfSchizont Extract IgG Seroprevalence (%) | 2.51 (3.93) | -5.20, 10.22 | 0.523 | 18 | 5 | [128-130, 134, 137] |
| log PvAMA1 IgG Seroprevalence (%) | 1.95 (0.08) | 1.79, 2.11 | <0.001 | 115 | 2 | [127, 134] |
| log PvMSP1<sub>19</sub> IgG Seroprevalence (%) | 1.25 (0.04) | 1.17, 1.32 | <0.001 | 103 | 2 | [127, 134] |

Effects for each exposure represent separate generalised multilevel modelling analyses estimating the association between the log of the seroprevalence of antimalarial antibodies and the seroprevalence of anti-gSG6 IgG, with the inclusion of a random intercept for study-specific heterogeneity and a random coefficient to allow the effect of the antimalarial antigen to vary across studies. Table shows log odds ratio and standard error (SE), 95% confidence interval (95%CI) and p-value, number of meta-observations (Meta-N) and studies, with associated references. Random effects not shown.
<sup>a</sup> Studies did not include a random coefficient (*i.e.* slope); as empirical support was not shown.

**Table S1:** Association between fSG6 IgG seropositivity and human biting rate

| Variable | Log Odds Ratio (SE) | 95% CI | p-value | RE |
| --- | --- | --- | --- | --- |
| **Fixed part** |  |  |  |  |
| log HBR | 0.17 (0.07) | 0.03, 0.31 | 0.017 |  |
| **Random part** |  |  |  |  |
| $\psi_1^c$ | | | | 0.47 |
| $\rho_1^d$ | | | | 0.13 |

Association between human biting rate (HBR) and fSG6 IgG: log odds ratio and standard error (SE), 95% confidence interval (95%CI), p-value, random-effect components (RE): variances ( $\psi$ ), conditional intraclass correlation coefficient ( $\rho$ )<sup>a</sup> and model log likelihood ( $\ell$ ) from generalised multilevel modelling.<sup>b</sup> This analysis is based upon n=6 meta-observations.
<sup>a</sup> $\rho = \frac{\psi_k + \dots + \psi_{nk}}{\psi_k + \dots + \psi_{nk} + \pi^2/3}$ , where $\psi_k$ through $\psi_{nk}$ are random-effect variance estimates pertaining to each of the respective variance components (see table notes <sup>c-d</sup>) from the mixed-effect generalised (logit) linear mixed models for a specific ICC estimate.
<sup>b</sup> Generalised multilevel modelling estimating the association between anti-*An. funestus* fSG6 IgG seropositivity and log transformed HBR with random-effects for study-specific heterogeneity in fSG6 IgG seropositivity.
<sup>c</sup> $\psi_1$ represents variance of the random-effect for study.
<sup>d</sup> $\rho_1$ represents conditional ICC for meta-observations from the same study.

**Table S2:** Association between gSG6-P2 IgG seropositivity and log PfCSP IgG seroprevalence

| Variable | Log Odds Ratio (SE) | 95% CI | p-value | RE |
| --- | --- | --- | --- | --- |
| **Fixed part** |  |  |  |  |
| log PfCSP IgG Seroprevalence | 3.70 (0.12) | 3.45, 3.94 | <0.001 |  |
| **Random part** |  |  |  |  |
| $\psi_1^c$ | | | | 25.2 |
| $\rho_1^d$ | | | | 0.88 |

Association between log PfCSP seroprevalence and gSG6-P2 IgG: log odds ratio and standard error (SE), 95% confidence interval (95%CI), p-value, random-effect variances ( $\psi$ ), conditional intraclass correlation coefficient ( $\rho$ )<sup>a</sup> and model log likelihood ( $\ell$ ) from generalised multilevel modelling.<sup>b</sup> This analysis is based upon n=115 meta-observations.
<sup>a</sup> $\rho = \frac{\psi_k + \dots + \psi_{nk}}{\psi_k + \dots + \psi_{nk} + \pi^2/3}$ , where $\psi_k$ through $\psi_{nk}$ are random-effect (RE) variance estimates pertaining to each of the respective variance components (see table notes <sup>c-d</sup>) from the mixed-effect generalised (logit) linear mixed models for a specific ICC estimate.
<sup>b</sup> Generalised multilevel modelling estimating the association between log PfCSP seroprevalence and anti-gSG6-P2 IgG seropositivity with random-effects for study-specific heterogeneity in gSG6-P2 IgG seropositivity.
<sup>c</sup> $\psi_1$ represents variance of the random-effect for study.
<sup>d</sup> $\rho_1$ represents conditional ICC for meta-observations from the same study.

**Table S3:** Association between gSG6-P2 IgG seropositivity and log PfGLURP IgG seroprevalence

| *Variable* | *Log Odds Ratio (SE)* | *95% CI* | *p-value* | *RE* |
| --- | --- | --- | --- | --- |
| *Fixed part* |  |  |  |  |
| log PfGLURP IgG Seroprevalence | 2.01 (0.09) | 1.85, 2.18 | <0.001 |  |
| *Random part* |  |  |  |  |
| $\psi_1^c$ | | | | 24.3 |
| $\rho_1^d$ | | | | 0.88 |

Association between log PfGLURP seroprevalence and gSG6-P2 IgG: log odds ratio and standard error (SE), 95% confidence interval (95%CI), p-value, random-effect variances ( $\psi$ ), conditional intraclass correlation coefficient ( $\rho$ )<sup>a</sup> and model log likelihood ( $\ell$ ) from generalised multilevel modelling.<sup>b</sup> This analysis is based upon n=116 meta-observations.
<sup>a</sup> $\rho = \frac{\psi_k + \dots + \psi_{nk}}{\psi_k + \dots + \psi_{nk} + \pi^2/3}$ , where $\psi_k$ through $\psi_{nk}$ are random-effect (RE) variance estimates pertaining to each of the respective variance components (see table notes <sup>c-d</sup>) from the mixed-effect generalised (logit) linear mixed models for a specific ICC estimate.
<sup>b</sup> Generalised multilevel modelling estimating the association between log PfGLURP seroprevalence and anti- gSG6-P2 IgG seropositivity with random-effects for study-specific heterogeneity in gSG6-P2 IgG seropositivity.
<sup>c</sup> $\psi_1$ represents variance of the random-effect for study.
<sup>d</sup> $\rho_1$ represents conditional ICC for meta-observations from the same study.

